# Social risk factors for SARS-CoV-2 acquisition in University students: cross sectional survey

**DOI:** 10.1101/2021.07.15.21260006

**Authors:** Eleanor Blakey, Lucy Reeve, Neville Q Verlander, David Edwards, David Wyllie, Mark Reacher

## Abstract

**Objectives:** To define risk factors for SARS-CoV-2 infection in University of Cambridge students during a period of increased incidence in October and November 2020.

**Study design:** Survey

**Methods:** Routine public health surveillance identified a marked increase in the numbers of University of Cambridge students with respiratory illness and SARS-CoV-2 positivity in the 10 days after a national lockdown was announced in the UK on 5 November 2020. Cases were identified both through symptom-triggered testing and a universal asymptomatic testing program. An online questionnaire was sent to all University of Cambridge students on 25 November to investigate risk factors for testing positive in the period after 30^th^ October 2020. This asked about symptoms, SARS-CoV-2 test results, in-person teaching settings, other aspects of University life, and attendance at social events in the period just prior to lockdown, from 30^th^ October and 4^th^ November 2020. Univariate and multivariable analyses were undertaken evaluating potential risk factors for SARS-CoV-2 positivity.

**Results:** Among 3,980 students responding to the questionnaire, 99 (2.5%) reported testing SARS-CoV-2 positive in the period studied; 28 (28%) were asymptomatic. We found strong independent associations with SARS-CoV-2 positivity were attendance at two social settings in the City of Cambridge (adjusted odds ratio favouring disease 13.0 (95% CI 6.2,26.9) and 14.2 (95% CI 2.9,70)), with weaker evidence of association with three further social settings. By contrast, we did not observe strong independent associations between disease risk and type of accommodation or attendance at, or participation in, a range of activities associated with the University curriculum.

**Conclusions:** Attendance at social settings can facilitate widespread SARS-CoV-2 transmission in University students. Constraint of transmission in higher education settings needs to emphasise risks outside University premises, as well as a COVID-safe environment within University premises.

**Highlights:**

- In a population of University students, a large increase in individuals testing positive for SARS-CoV-2 occurred in the days following a national lockdown.
- Attendance at particular social gatherings was strongly linked to the development of disease, independent of other risk factors.
- By contrast, a range of risk factors including age, gender, ethnicity, accommodation type, shared kitchen facilities, attendance at supermarkets, and attending teaching sessions were not significantly associated with SARS-CoV-2 risk.
- These data emphasise the increased risk associated with University students attending social settings with large numbers of others, even when other risks associated with university attendance are well controlled.

## Introduction

Universities have been identified as sites where SARS-CoV-2 transmission can readily occur. Along with other sectors of the economy, social distancing and other non-pharmaceutical interventions were mandated by the UK government in higher education during the coronavirus pandemic[1]. These measures were informed by disease transmission modelling[2] and by experiences gained earlier in the pandemic[3].

The University of Cambridge used the guidelines outlined by the UK government to create its own set of guidelines that could be implemented in its 31 constituent colleges. Each of these colleges range in geographical size, and in the demographic of their annual intakes. It is in these colleges that students reside, undertake most if not all of their in-person teaching, and attend college-led formal and informal social events.

At the point the University of Cambridge student year began in October 2020[4], the B.1.1.7 SARS-CoV-2 variant was transmitting widely in parts of the UK[5]. Students returning to Universities found themselves studying in ‘COVID-safe’ environments featuring many changes on the pre-SARS-CoV-2 regime, included altered housing arrangements with students mixing in small groups (‘bubbles’), the wearing of masks, social distancing during tuition, and the use of distance learning approaches. In addition, some universities put in place free voluntary PCR-based screening programs for students; the UoC was one, offering an asymptomatic screening programme which is described elsewhere[6]. Such programs complemented the provision of PCR testing for symptomatic individuals by the state, free of charge at the point of use.

Despite the control measures described, outbreaks occurred among students across the UK Higher education sector, which includes over 2.3 million students in 160 institutions. The determinants of these outbreaks are still being studied; residence in larger halls of residence has been identified as one risk factor[3], but determinants of successful COVID-safety in higher education settings are still unclear.

We address this by analysing risk factors for SARS-CoV-2 acquisition in one UK university in Cambridge in the period prior to England’s second national lockdown (which commenced 5^th^ November 2020). Up to this time, rates among individuals aged under 60 years were generally increasing across England[7] but in Cambridge local authority these case rates were stable[8]. Public Health England, a statutory body tasked with outbreak surveillance, became aware of increased incidence in a number of University of Cambridge (UoC) Colleges, identified through both the national symptom-derived SARS-CoV-2 testing and through the work of the University of Cambridge asymptomatic screening programme which screened asymptomatic students weekly[6, 9]. PHE conducted an analytical epidemiological survey into the determinants of SARS-CoV-2 incidence, results of which we present and discuss here.

## Methods

### Participants

The study population targeted were University of Cambridge (UoC) students resident in Cambridge during the study period. This was a subset of the 25,256 UoC students who would reside in Cambridge City during Michaelmas term 2020 under normal circumstances. However, the pandemic situation meant not all students in the study population were residing in Cambridge at that time. We identified the study population from the 25,256 student group as part of a questionnaire sent to all students: residency was determined through a student’s response to the initial survey question. Students who answered as not residing in Cambridge City during this period were taken to the finishing page of the survey and excluded from analytical epidemiological studies.

### Survey

Cases were defined as individuals testing positive for SARS-CoV-2, with or without symptoms, between 30^th^ October 2020 and the date of questionnaire completion, which was 25^th^ or 26^th^ November in almost all cases (see below). All other respondents were considered controls; this included individuals with SARS-CoV-2 like symptoms who either had no test result or a non-positive test result.

Risk factors assessed by the questionnaire administered (included in Supplementary Materials) included: age, gender, ethnicity, UoC college, student type (undergraduate or postgraduate), symptoms of COVID-19 and SARS-CoV-2 test results, term time accommodation, food shopping habits, travel habits, and in-person teaching settings. We also asked about attendance, queueing and social distancing at social events attended between Friday 30^th^ October and Wednesday 4^th^ November 2020. We focused on these exposures because of anecdotal observations by colleagues that a number of affected students may have visited such venues.

It was decided to conduct the questionnaire on a de-identified (unnamed) basis in order to encourage full and honest responses from students. Thus, results of tests are self-reported. While questionnaires asked about attendance at a defined set of named social settings and venues for event attendance, the identities of which have been anonymised as ‘social setting’ or ‘SS’ followed by a number.

The study population were contacted by email with a link to the online questionnaire, which was hosted in Snap Survey, a commercial questionnaire software, on 25^th^ November 2020. The questionnaire was live for one week before it closed to responses at 12:00pm on 3^rd^ December 2020.

### Data analysis

#### Descriptive analysis

Age, gender, ethnicity, UoC college and symptoms of cases and non-cases were described.

#### Data cleaning

Prior to inferential analysis, we generated numerical fields created from open-ended text fields describing other exposures not listed in the questionnaire whereby those individuals not explicitly mentioning the exposure had their values changed from missing to ‘No’. This applied to the fields of ‘college catering’, ‘attended labs’, ‘attended seminar(s)’, ‘medical student placement’ and ‘met in other accommodation’. All other data entries were analysed as entered.

#### Mixed effects logistic regression

was used with the binary response (SARS-CoV-2 positive/negative) as the outcome and student college as the random effect to allow for possible non-independence between student outcomes. The odds ratio (OR) as the measure of effect was used and it, together with its 95% confidence interval (CI), are quoted in the results. The p-values were obtained by means of the likelihood ratio test or, if not possible, the Wald test. A statistical significance level of p≤0.05 was chosen.

The analysis began by conducting a univariate analysis. This involved fitting a series of models, each with just one fixed effect without regards to other explanatory variable and considering each factor in turn. Those variables with p-value of 0.2 or less, odds ratio larger than 1.0 and the variables of queueing and social distancing at events were then considered further in a multivariable model in a backwards stepwise procedure wherein, at each step, a fixed effect with the most missing values among those not considered by that stage and p-value larger than 0.1 was considered for removal from the model. It was removed if it was not substantially confounding. A variable was considered to be substantially confounding if its removal resulted in a change of 10% or more in one or more of the odds ratios for the variables still in the model. The process concluded with the final model when each of the variables in the model met one or more of the following: had been found to be substantially confounding in the one of the preceding steps, had a p-value of 0.1 or less, or removal would not increase the number of available observations with which to perform a complete-case analysis. The adequacy of excluding the variables dropped during the model building process was checked by adding them one at a time to the final model (and removing it before adding another) to see that each remained non-significant and was not substantially confounding.

For the continuous variable in the dataset (age), a stepwise procedure was performed by beginning with a cubic function (on the logit scale) and simplifying to the next simplest function if the deterioration in fit was not statistically significant until either the function was linear or the least complex function not fitting significantly worse. This was done in the single variable analysis as well as the first step in the multivariable modelling procedure. After this first step, implausible protective factors were removed one at a time until there were no such fixed effects. The subsequent multivariable modelling steps followed the process described above.

All analyses were performed in Stata (StataCorp) versions 15 and 16.1.

## Results

### Cohort studied

The online questionnaire was deployed to 25,256 UoC students. In total 4,447 questionnaires were returned, which contained 1,151 incomplete responses giving a response rate of 17.6%. We excluded 78 ineligible responses from individuals other than students, and 389 incomplete responses without details of symptoms or key demographic data, leaving 3,980 responses in the final analysis (15.8% analysable rate) (Figure 1).

**Figure 1:**
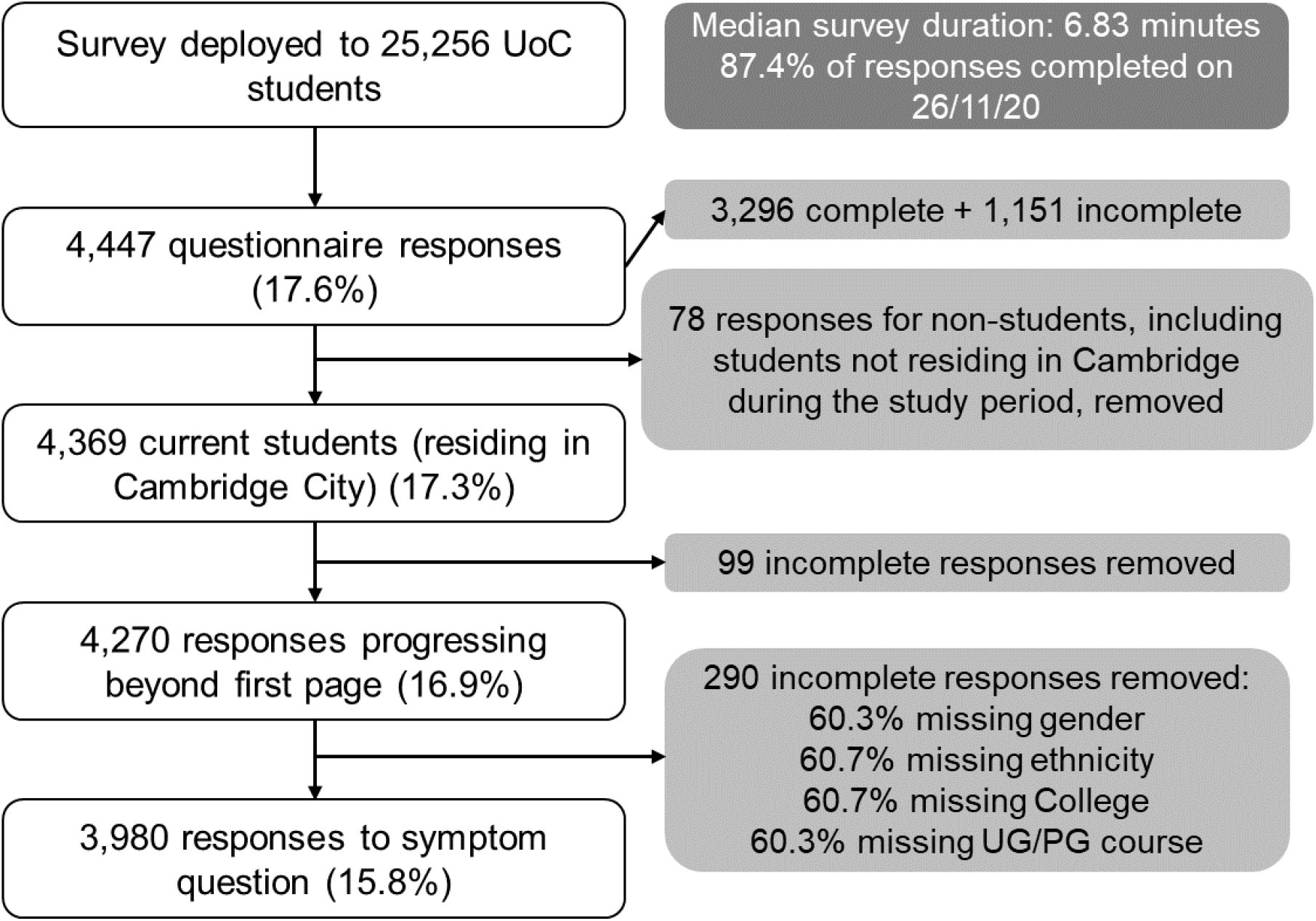
Questionnaire responses, University of Cambridge cohort.

Out of the 3,980 responses used in the final analysis, 99 individuals met the case definition (2.5%) of a positive individual test result for SARS-CoV-2 on or after 30^th^ October 2020, while 3,617 individuals did not (264 individuals were unable to be categorised as either – they did not answer any specimen questions). As expected, the reported durations of illness in these individuals matched a spike in incidence reported in Cambridge local authority between 8^th^ and 12^th^ November 2021[8], detected by national surveillance systems (see supplementary Figure 1).

Responses, and positive cases, were received from across Cambridge’s colleges (Figure 2, Appendix 1). The demographic details of respondents is typical of Cambridge students (Figure 3); the median age was 20 years, with respondents being predominantly white (2,935, 74%), while 60% (2,386) were undergraduate students (2,386) (Table 1, Figure 3). For more demographic details, see Appendix 2.

**Table 1:**
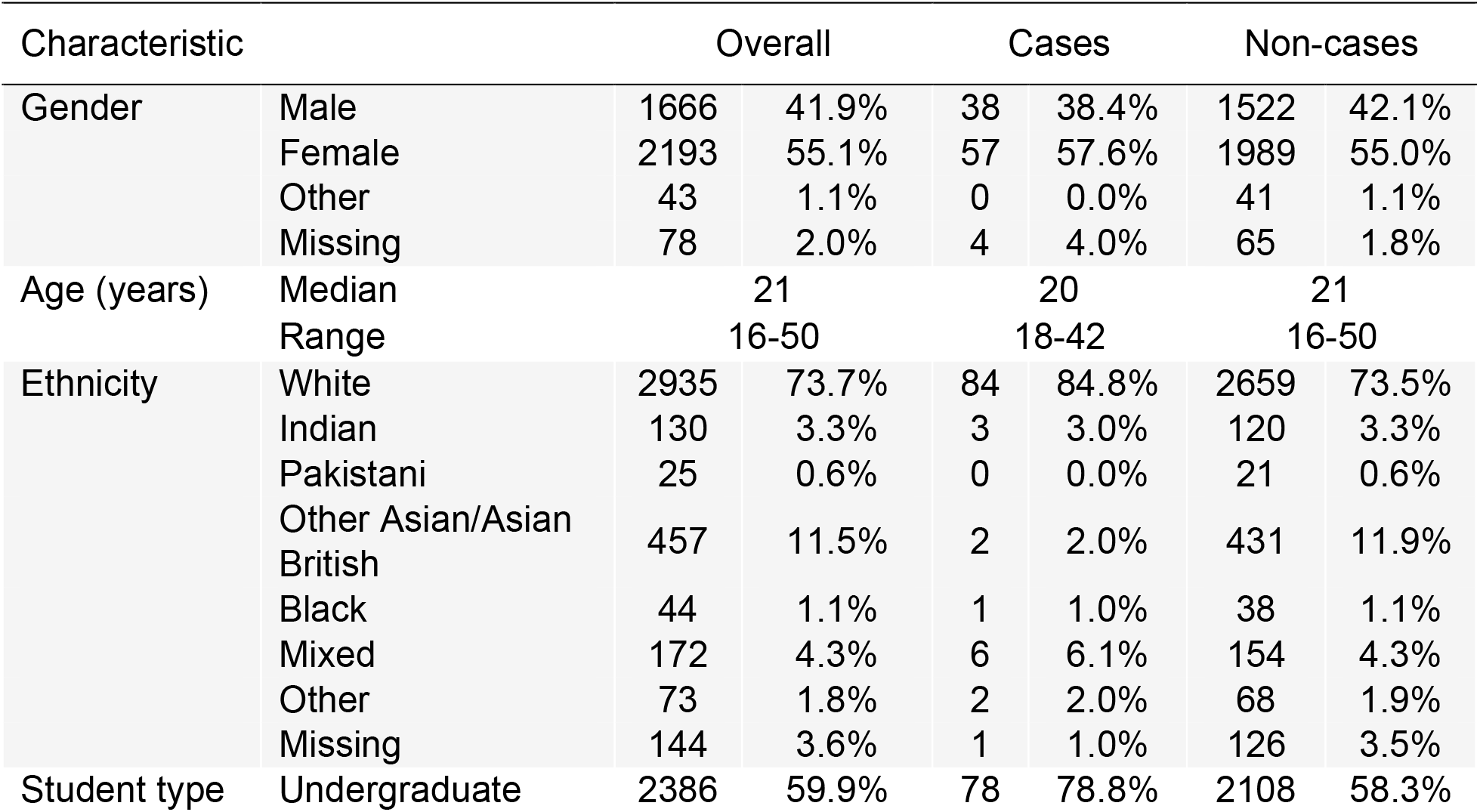
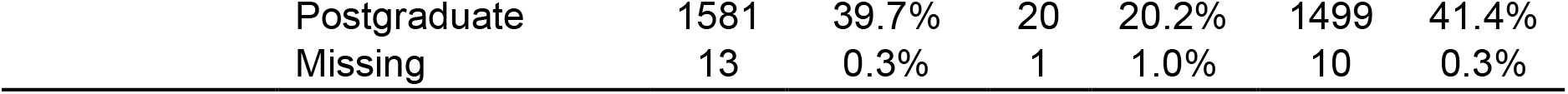
Characteristics of cases and non-cases, University of Cambridge cohort (n=3,980)

**Figure 2:**
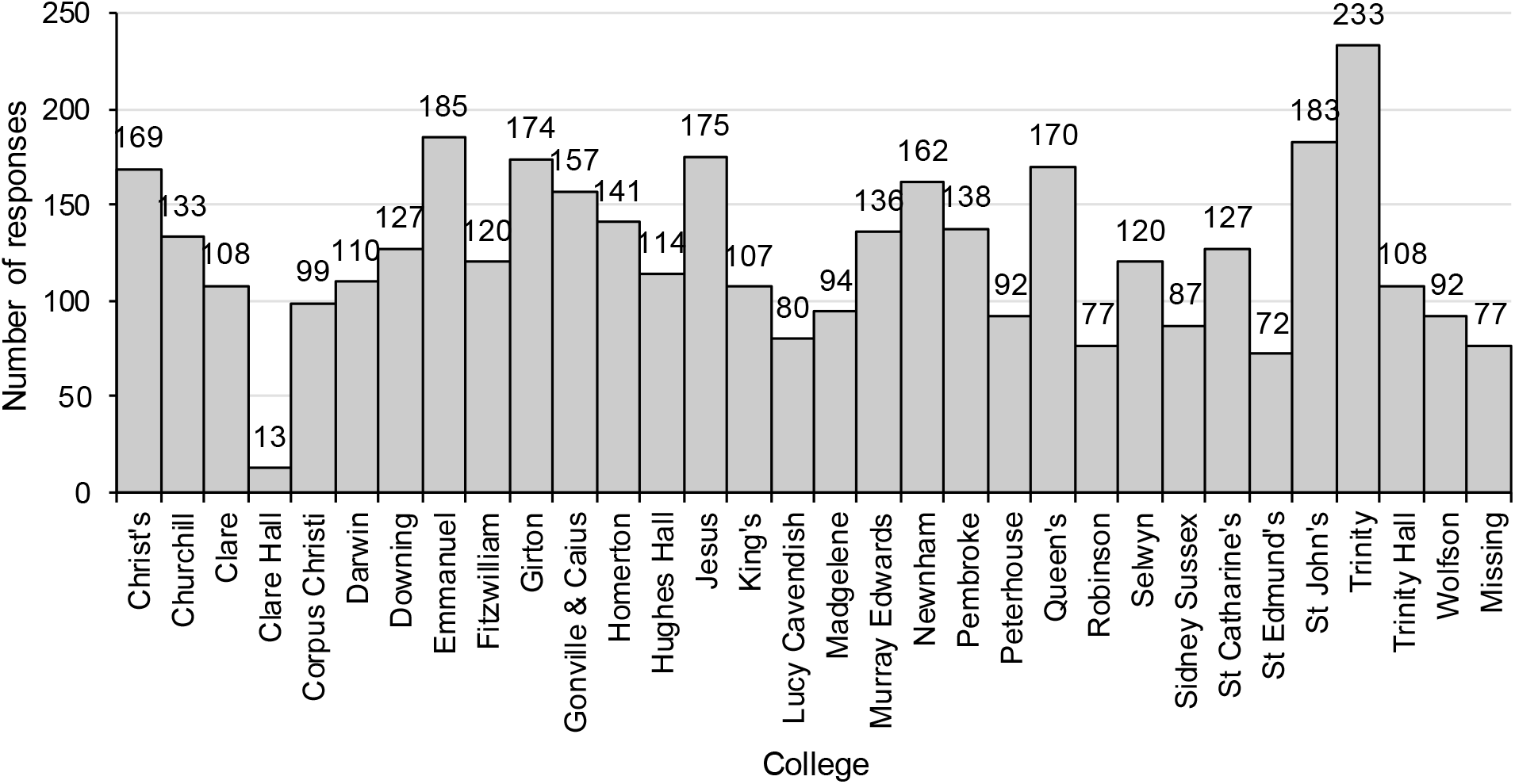
Distribution of cases and non-cases among University of Cambridge colleges (n=3,980)

**Figure 3:**
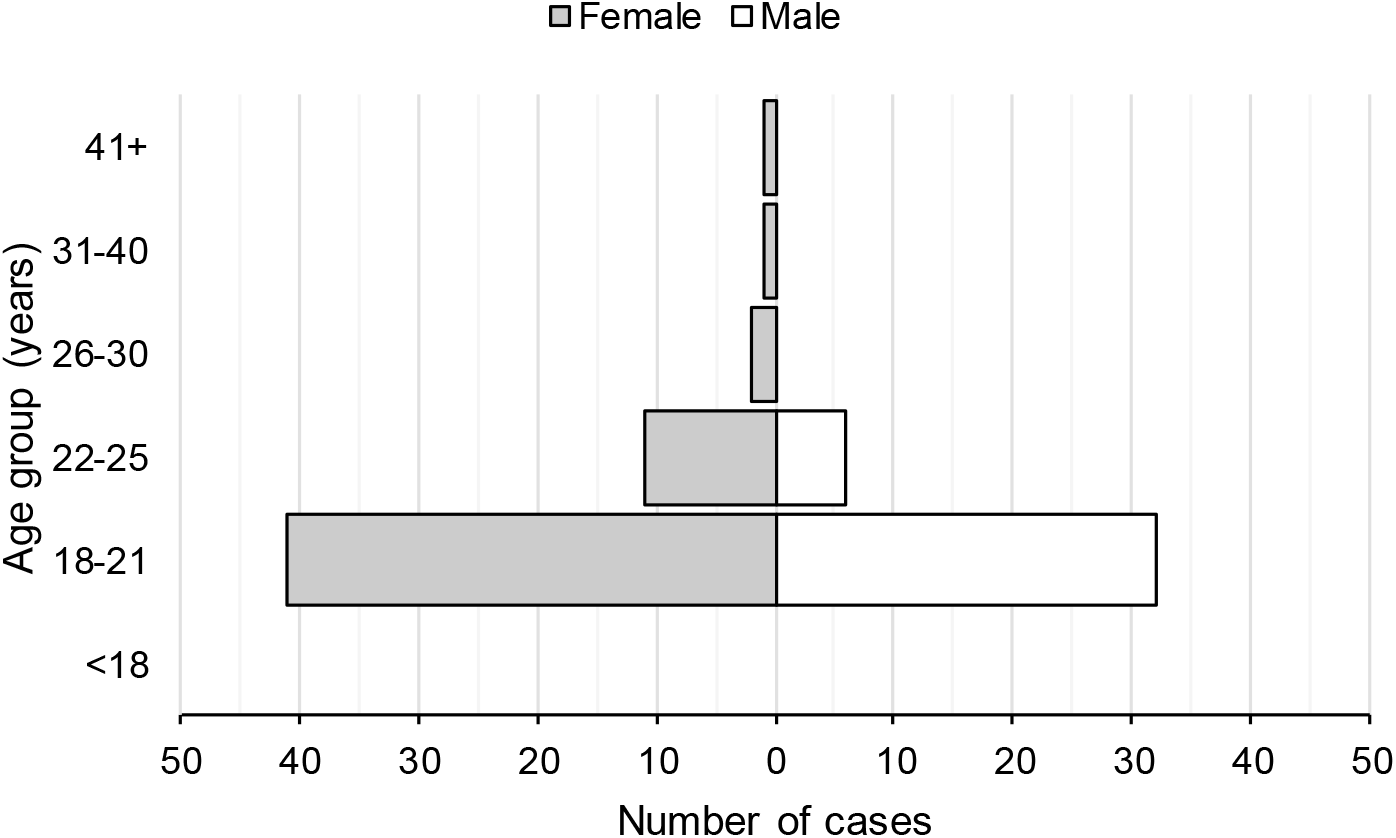
Age and gender distribution among cases, University of Cambridge cohort (n=94)

**Figure 3:**
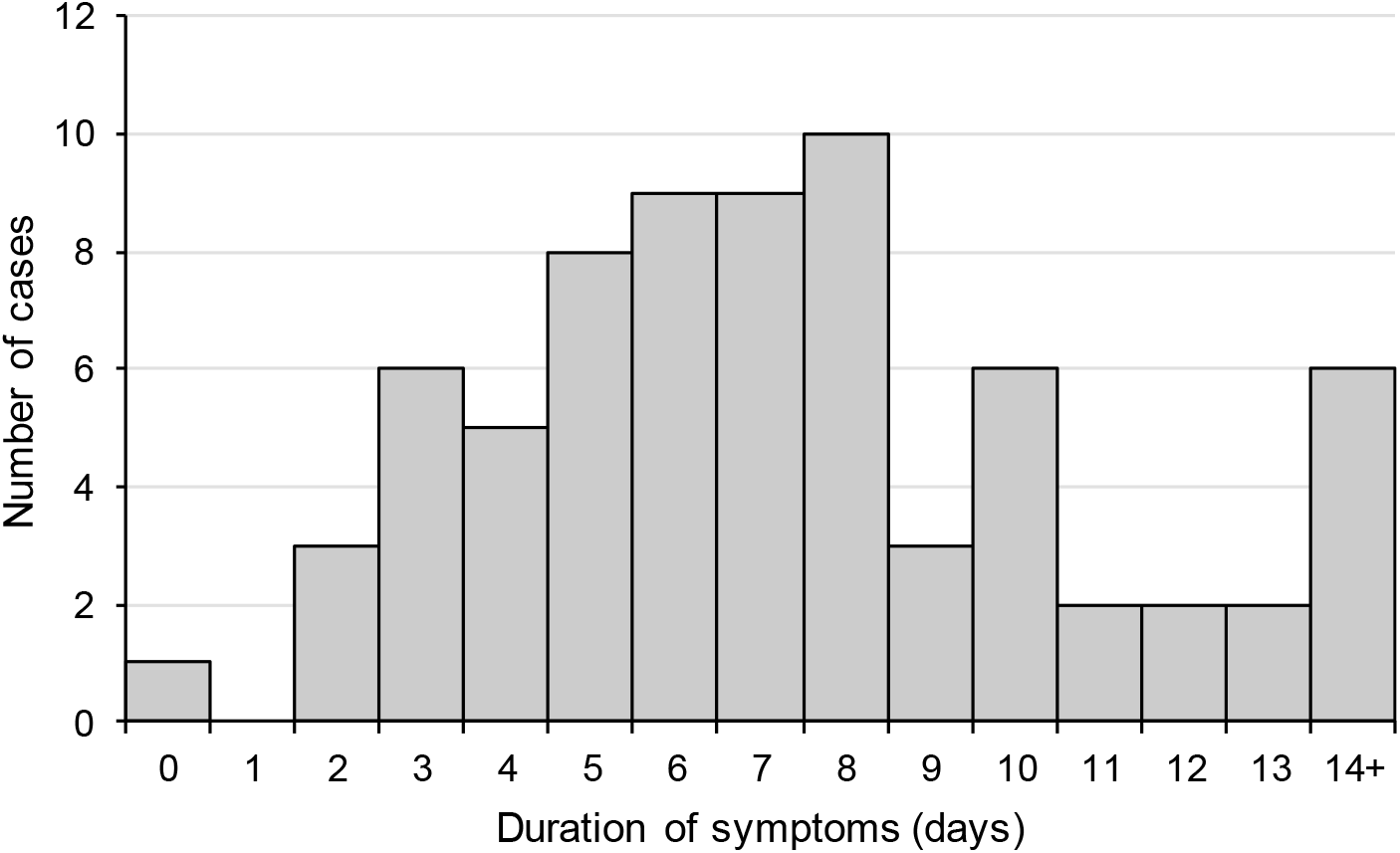
Distribution of the duration of symptoms among cases, University of Cambridge cohort (n=72)

### Individuals testing positive

Ninety-nine individuals reported testing positive. The majority of cases reported that their positive result was part of the University of Cambridge screening programme (66.7%), with smaller proportions detected by NHS testing (21.2%) and the Cambridge University Hospital screening programme (11.1%). Nearly half of cases (45.5%) reported having had face to face contact with another known case of COVID-19 since 16th October 2020, compared to 9.8% of non-cases.

The earliest date of symptom onset was 27^th^ October, with the majority of cases reporting onset after the start of national lockdown on 5^th^ November, peaking at 15 cases with onset on 10^th^ November (Figure 4). Table 2 shows the frequency of symptoms reported among cases, of which the most commonly reported symptoms were COVID-like illness (77.8%) of fever or cough or loss/change of sense of taste/smell, headache (76.8%), sore throat (67.7%), fatigue (64.6%) and runny nose (61.6%). More than one quarter of cases were asymptomatic (28.3%). The median duration of illness was 7 days, ranging from 0 to 38 days, with the full distribution shown in Figure 5. The majority of cases (60) recovered within ten days of symptom onset. For all symptoms except vomiting, the frequency of self-reported symptoms in cases was much higher than in non-cases (Table 2). Details of healthcare consultations following COVID-19 diagnosis are in Table 3.

**Table 2:**
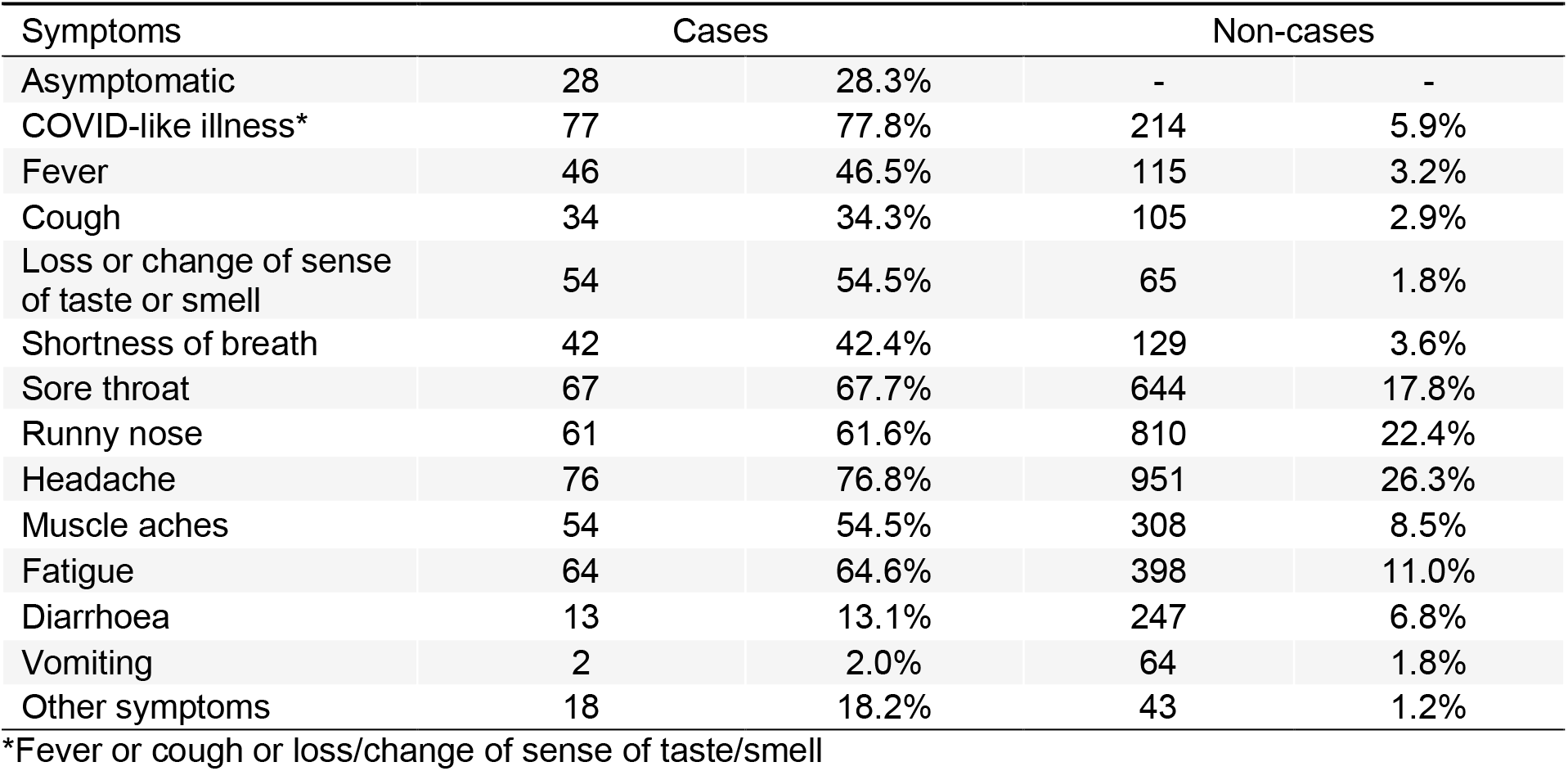
Frequency of symptoms among cases and non-cases, University of Cambridge cohort (n=3,980)

**Table 3:** Type of healthcare consulted by cases, University of Cambridge cohort (n=91)

| Type of healthcare consulted | Cases |  |
| --- | --- | --- |
| College nurse | 17 | 18.7% |
| NHS 111 | 5 | 5.5% |
| GP or another doctor | 3 | 3.3% |
| Visited A&E | 0 | 0.0% |
| Other healthcare | 2 | 2.2% |
| Admitted to hospital | 0 | 0.0% |

**Figure 4:**
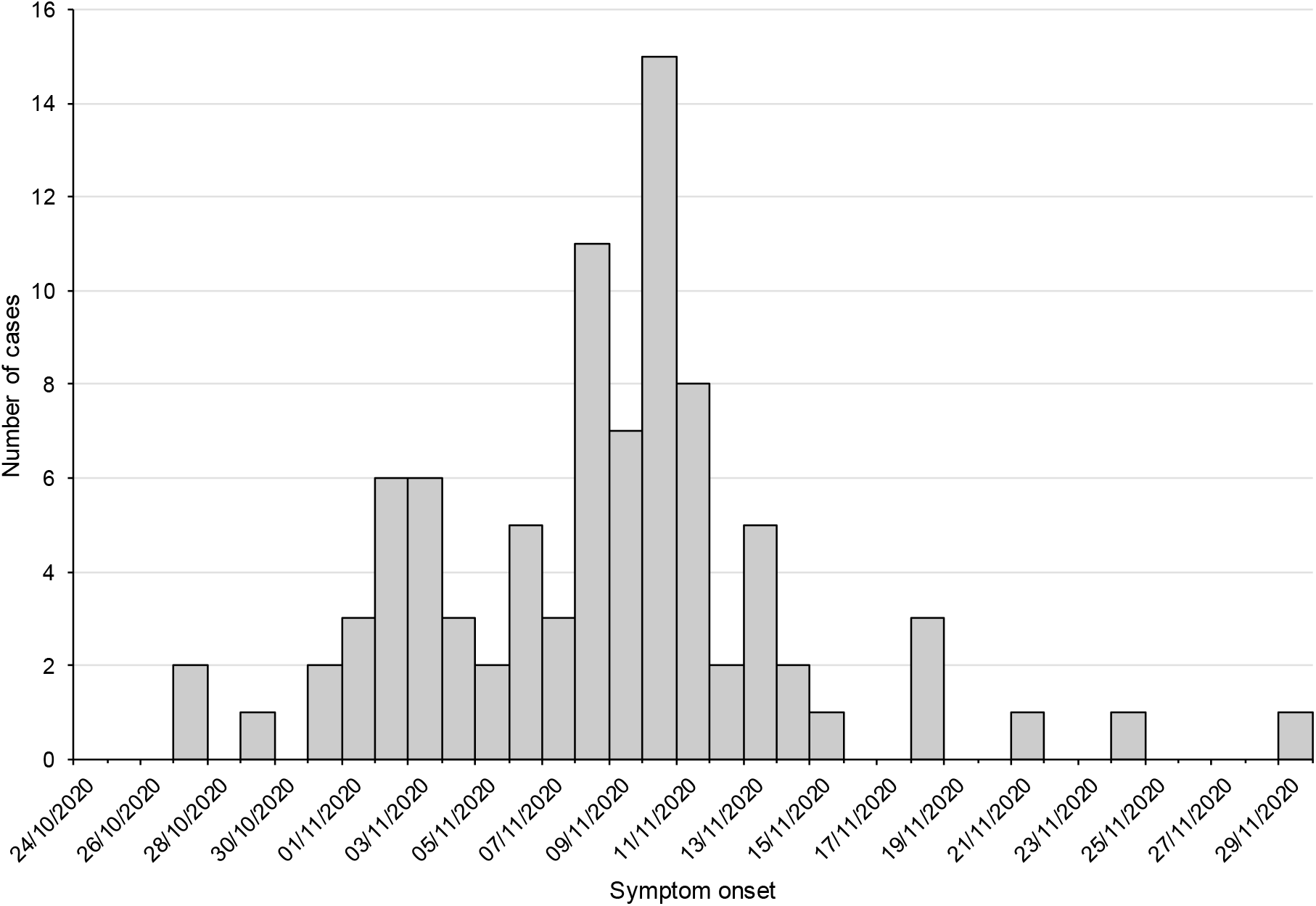
Distribution of cases by date of symptom onset reported, University of Cambridge cohort (n=90)

### Risk factors for SARS-CoV-2 test positivity

In univariate analyses, we observed strong associations between testing positive and attendance at some social settings (most notably attendance at SS7 or SS23). An association with SS3 (representing attendance at Formal Hall) was also noted (OR 2.73, 95% CI 1.2, 6.4). Depending on the college, formal hall is an all college weekly tradition where attendees dine together in a communal dining hall. Attendees are seated by academic rank with senior academics dining at a ‘high’ or separated table. Other University associated activities (Appendix 3, Teaching section, as well as Social Settings 1,4,6 and 35, which were University related) were not significantly associated with disease. In univariate analyses, we also noted disease association with being a postgraduate vs. undergraduate, and with type of accommodation (see Table 4 for variables subsequently included the multivariable analysis, and Appendix 3 for all other exposures).

**Table 4:**
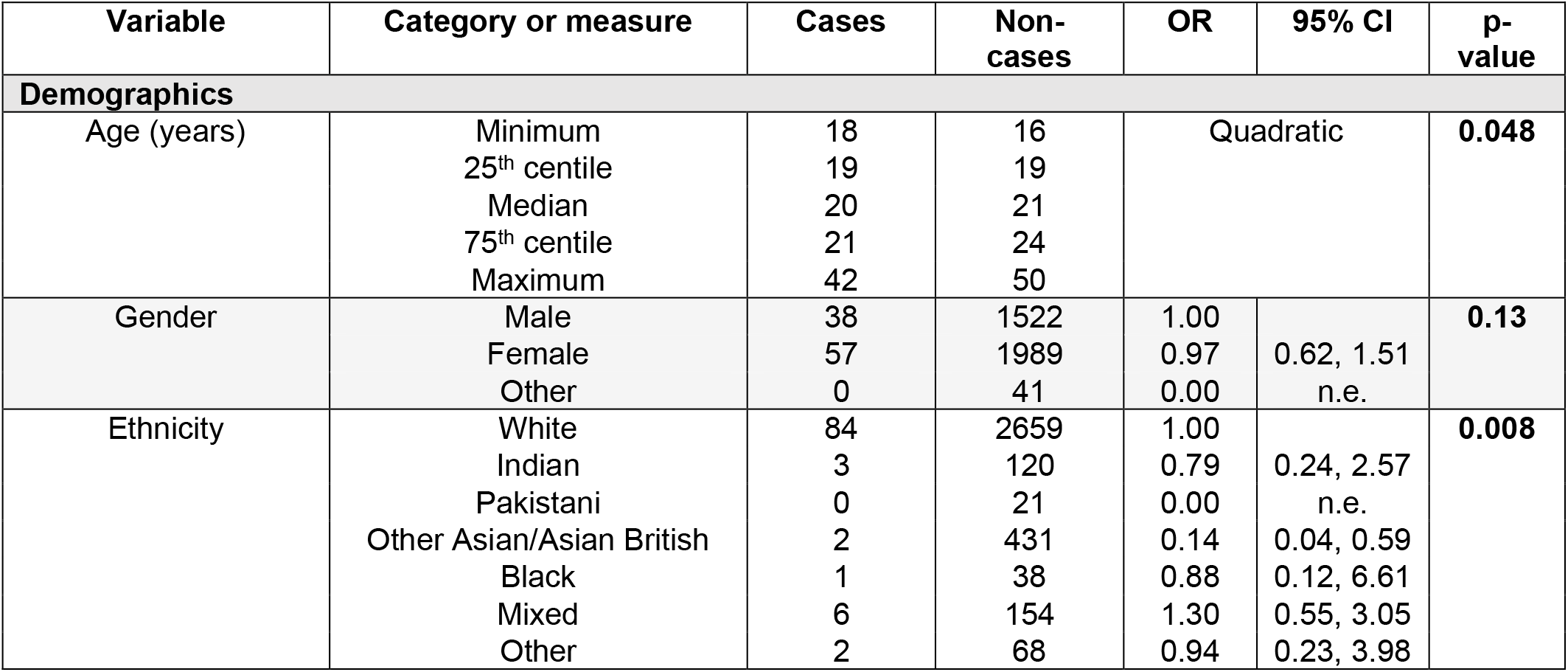
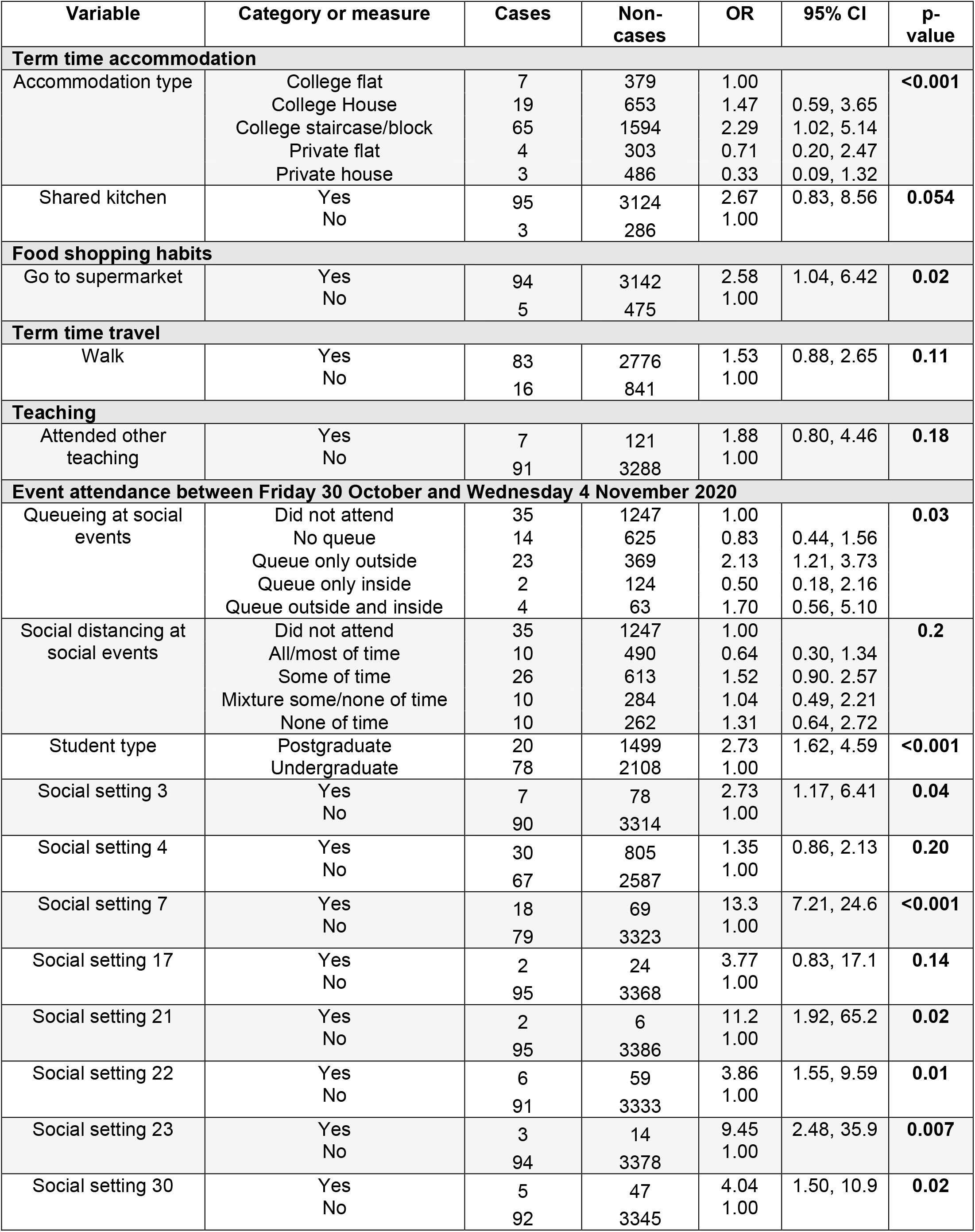
Single variable analysis of demographics, lifestyle and social event exposures among cases and non-cases, University of Cambridge cohort.

### Multivariable Analysis

Following univariate analysis, nineteen variables were considered in the multivariable model (see Methods for selection criteria, and Table 5). We removed terms making minimal contributions to the model (see Methods), eliminating the terms ‘go to supermarket’, ‘shared kitchen’, ‘gender’, ‘other teaching’, ‘social setting (SS) 4’ ‘SS21’, ‘SS22’, ‘walking’, and ‘student type’. We also removed the variable on Queuing at social events. We did this because it is possible that outdoor queueing is a marker of COVID-19 safe environments to which access is restricted, making interpretation difficult without stratification by venue, something we explore further below. The final model is shown in Table 6.

**Table 5:** Multivariable model (n=2252), University of Cambridge cohort.

| Variable | Category or measure | OR | 95% CI | p-value |
| --- | --- | --- | --- | --- |
| Age | Per year | 0.94 | 0.84, 1.04 | 0.5 |
| Ethnic group | White | 1.00 |  | 0.02 |
|  | Indian | 1.01 | 0.23, 4.51 |  |
|  | Pakistani | 0.00 | n.e. |  |
|  | Other Asian/Asian British | 0.10 | 0.01, 0.77 |  |
|  | Black | 0.00 | n.e. |  |
|  | Mixed | 1.34 | 0.44, 4.06 |  |
|  | Other | 1.09 | 0.14, 8.61 |  |
| Accommodation type | College flat | 1.00 |  | 0.6 |
|  | College House | 1.11 | 0.40, 3.11 |  |
|  | College staircase/block | 1.29 | 0.50, 3.30 |  |
|  | Private flat | 0.69 | 0.16, 3.01 |  |
|  | Private house | 0.35 | 0.07, 1.91 |  |
| Queueing at events | Not attended | 1.00 |  | 0.8 |
|  | No queueing | 0.71 | 0.26, 1.97 |  |
|  | Queueing only outside | 0.60 | 0.20, 1.83 |  |
|  | Queueing only inside | 0.38 | 0.07, 2.17 |  |
|  | Queueing inside and outside | 0.41 | 0.09, 1.89 |  |
| Social distancing (SD) at events | Not attended | 1.00 |  | 0.15 |
|  | SD all/most of time | 0.40 | 0.12, 1.39 |  |
|  | SD some of time | 1.17 | 0.45, 3.08 |  |
|  | Mixture of SD some/none of time | 0.57 | 0.17, 1.87 |  |
|  | SD none of time | n.e. | n.e. |  |
| Social setting 3 | Yes | 3.03 | 0.96, 9.53 | 0.08 |
|  | No | 1.00 |  |  |
| Social setting 7 | Yes | 13.9 | 5.52, 57.2 | <0.001 |
|  | No | 1.00 |  |  |
| Social setting 17 | Yes | 10.0 | 1.76, 57.2 | 0.09 |
|  | No | 1.00 |  |  |
| Social setting 23 | Yes | 17.3 | 3.01, 99.4 | 0.004 |
|  | No | 1.00 |  |  |
| Social setting 30 | Yes | 3.81 | 1.02, 14.2 | 0.06 |
|  | No | 1.00 |  |  |

**Table 6:**
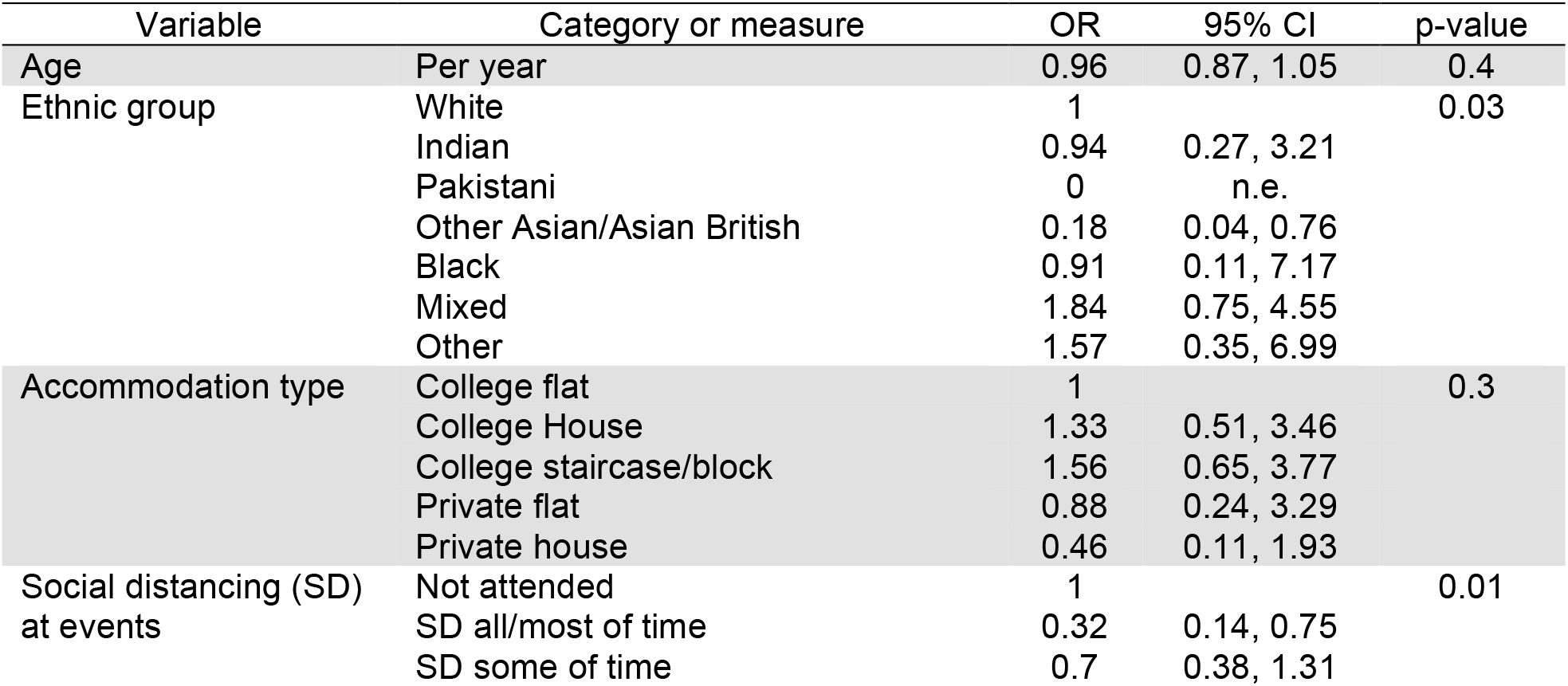
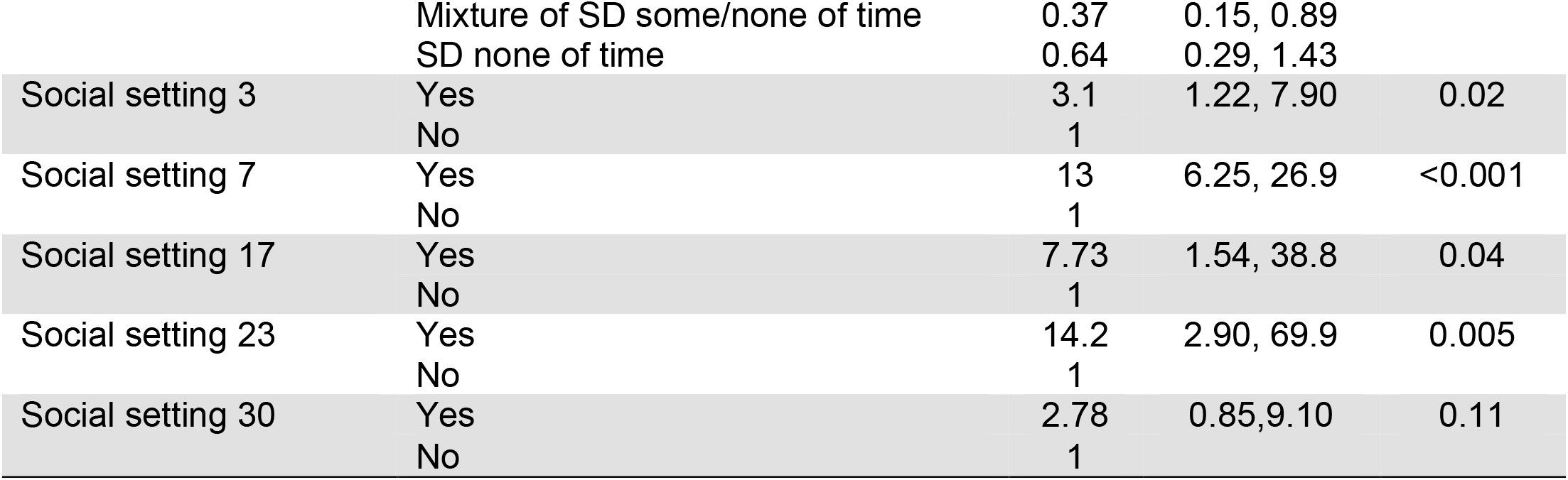
Final multivariable model without “queueing at social events” (n=2825), University of Cambridge cohort.

In the final multivariable model (Table 6) and univariable analysis (Table 5) both the strongest independent associations with positivity were attendance at SS7 (unadjusted OR 13.9 (95% CI: 5.52, 57.2); adjusted OR (aOR) 13.0 (6.25, 26.9)) and SS23 (OR 17.3 (95% CI: 3.01, 99.4); aOR 14.2 (2.90, 69.9)). There is also some evidence of independent association with positivity, of attendance at social settings SS3 (Formal Hall), and with SS17 and SS30, which are commercial venues at which socialisation occurred. The strength of the observed effect differed slightly depending on the model used (Table 5 vs. Table 6).

Interestingly, of the students that attended SS7 and answered questions about queuing and social distancing (n=68), there were variable reports about the extent of social distancing, with 33 respondents stating that social distancing was practiced all the time (33 responses) and 39 responding that it was practiced some of the time. In SS23, 17 of the respondents reported social distancing being practiced all (8 responses), or some of the time (5 responses). See Appendix 4 for full tabulation.

It is notable that neither undergraduate/postgraduate status, nor attendance at University teaching form part of the final model, and that the contribution of accommodation type is not significant. It appears that the associations of SARS-CoV2 acquisition with these risk factors is captured by other risk factors, notably the attendance at social events.

## Discussion

This investigation has found strong evidence of independent association with SARS-CoV-2 detection and attendance at two social venues, with weaker evidence at others. The highest odds was with attendance at SS7 and SS23 (aOR 13.9 (95% CI 5.5,57) and aOR 17.3 (3,99) respectively, both were primarily settings where food and drink were served and consumed indoors. This is also a feature of SS3 (Formal Hall) which was similarly, but more weakly, associated with odds of disease (aOR 3.0, 95% CI 0.96, 9.5). Such indoor settings are recognised to represent a risk[10, 11]. In the time period of this study the wearing of face coverings in certain indoors setting was a mandatory legal requirement[12] unless sat at an assigned table. However, it would be expected, due to the consumption of food and beverages, that constant wearing of a face covering or mask would be difficult to maintain; adding to this point, respondents indicated inconstant social distancing. In contrast, it was notable that neither university attendance, type of residence, or student category contributed significantly to odds of positivity, and neither did attendance at the majority of university organised or based settings (SS1, SS2, SS4, SS6, and SS35). Responses received, and the timing of illness, suggests that socialising between students in non-university settings occurred shortly before a national lockdown was imposed, and in the context of a rapidly spreading SARS-CoV-2 epidemic. Our work also suggests that the control measures put in place by the university[13] were largely effective at minimising the odds of infection, with the possible exception of Formal Hall related dining.

This research study has several notable limitations. Firstly, by design it was anonymous, and so the responses obtained could not be checked against national information systems. Secondly, the survey was retrospective, so attendance at events or recording of symptoms may be subject to recall bias. Finally, we received 3,980 responses to the questionnaire from a potential study population of 25,256, a response rate of 15.8%. While there is evidence of external validity of the responses obtained – in particular, the time course of the development of illness reported matches what actually happened – the low response rates mean that the conclusions require external validation. Such external validation has recently been published, in the form of a genomic analysis of sequences from individuals with SARS-CoV-2 infection[9], which also strongly implicates social mixing outside of university settings as a key risk factor for SARS-CoV-2 infection. Further external validation can be seen through comparison of the cases reported in Cambridge local authority[8], and those by specimen date reported by the University of Cambridge cohort (see supplementary Figure 1). The cases in both figures peak around the same time period (8^th^ – 12^th^ November 2021).

This work builds on studies from elsewhere identifying indoor social settings as sites of SARS-CoV-2 transmission[11]. For example, at the start of the pandemic, nightclubs in Seoul reported multiple cases associated with venues of this type[14], and since the easing of social distancing and lockdown measures in South Korea, nightclubs have been highlighted as venues of concern, where cases could easily spread to the wider community[15]. Our results indicate that, in University settings, infection control measures aimed at establishing a low transmission ‘COVID-safe’ learning environment can readily be compromised by attendance at social gatherings.

### Ethics

This protocol was subject to a review by the PHE Research Ethics and Governance Group. It was classified as an outbreak investigation undertaken as part of PHE’s responsibility to respond to the COVID-19 current pandemic, and to inform the multi-agency response to the large rise in cases and future response. The study is anonymous and consent was requested to publish non-identifiable aggregate information derived from the study.

As such this work fell outside the remit for ethical review and as no regulatory issues were identified the protocol was approved.

## Supporting information

Supplementary: Survey

## Data Availability

An anonymised dataset supporting the findings of this study is available from the corresponding author on request.

## Acknowledgements

We thank the members of the University of Cambridge Outbreak Management Team, for their support and contribution to facilitating the questionnaire dissemination: Gillian Weale, Ian Jones and Robert Henderson from The University of Cambridge; Linda Sheridan, and Vickie Braithwaite from Cambridgeshire County Council; Yvonne O’Donnell from Cambridge City Council; Heidi Eagle from the East of England PHE East Health Protection Team; and The University of Cambridge communications team.

Appendices

### Appendix 1: Distribution of questionnaire responses by University of Cambridge college

| University of Cambridge college | Overall |  | Cases |  | Non-cases |  |
| --- | --- | --- | --- | --- | --- | --- |
| Christ's | 169 | 4.2% | 8 | 8.1% | 150 | 4.1% |
| Churchill | 133 | 3.3% | 2 | 2.0% | 124 | 3.4% |
| Clare | 108 | 2.7% | 3 | 3.0% | 101 | 2.8% |
| Clare Hall | 13 | 0.3% | 0 | 0.0% | 13 | 0.4% |
| Corpus Christi | 99 | 2.5% | 0 | 0.0% | 96 | 2.7% |
| Darwin | 110 | 2.8% | 1 | 1.0% | 103 | 2.8% |
| Downing | 127 | 3.2% | 2 | 2.0% | 116 | 3.2% |
| Emmanuel | 185 | 4.6% | 15 | 15.2% | 160 | 4.4% |
| Fitzwilliam | 120 | 3.0% | 6 | 6.1% | 104 | 2.9% |
| Girton | 174 | 4.4% | 7 | 7.1% | 154 | 4.3% |
| Gonville & Caius | 157 | 3.9% | 1 | 1.0% | 143 | 4.0% |
| Homerton | 141 | 3.5% | 1 | 1.0% | 131 | 3.6% |
| Hughes Hall | 114 | 2.9% | 2 | 2.0% | 106 | 2.9% |
| Jesus | 175 | 4.4% | 4 | 4.0% | 158 | 4.4% |
| King's | 107 | 2.7% | 1 | 1.0% | 90 | 2.5% |
| Lucy Cavendish | 80 | 2.0% | 3 | 3.0% | 71 | 2.0% |
| Madgelene | 94 | 2.4% | 3 | 3.0% | 85 | 2.4% |
| Murray Edwards | 136 | 3.4% | 13 | 13.1% | 115 | 3.2% |
| Newnham | 162 | 4.1% | 4 | 4.0% | 147 | 4.1% |
| Pembroke | 138 | 3.5% | 2 | 2.0% | 124 | 3.4% |
| Peterhouse | 92 | 2.3% | 0 | 0.0% | 85 | 2.4% |
| Queen's | 170 | 4.3% | 4 | 4.0% | 152 | 4.2% |
| Robinson | 77 | 1.9% | 2 | 2.0% | 70 | 1.9% |
| Selwyn | 120 | 3.0% | 2 | 2.0% | 109 | 3.0% |
| Sidney Sussex | 87 | 2.2% | 1 | 1.0% | 77 | 2.1% |
| St Catharine's | 127 | 3.2% | 2 | 2.0% | 114 | 3.2% |
| St Edmund's | 72 | 1.8% | 0 | 0.0% | 70 | 1.9% |
| St John's | 183 | 4.6% | 4 | 4.0% | 175 | 4.8% |
| Trinity | 233 | 5.9% | 2 | 2.0% | 220 | 6.1% |
| Trinity Hall | 108 | 2.7% | 0 | 0.0% | 98 | 2.7% |
| Wolfson | 92 | 2.3% | 1 | 1.0% | 91 | 2.5% |
| Missing | 77 | 1.9% | 3 | 3.0% | 65 | 1.8% |

### Appendix 2: Characteristics of responses with incomplete symptom reporting (n=290)

| Characteristic |  | Total | % |
| --- | --- | --- | --- |
| Sex | Male | 49 | 16.9% |
|  | Female | 66 | 22.8% |
|  | Missing | 175 | 60.3% |
| Ethnicity | White | 83 | 28.6% |
|  | Indian | 6 | 2.07% |
|  | Pakistani | 2 | 0.690% |
|  | Other Asian/Asian British | 17 | 5.86% |
|  | Black | 2 | 0.690% |
|  | Mixed | 3 | 1.03% |
|  | Other | 1 | 0.345% |
|  | Missing | 176 | 60.7% |
| College | Christ's | 6 | 2.07% |
|  | Churchill | 5 | 1.72% |
|  | Clare | 4 | 1.38% |
|  | Corpus Christi | 2 | 0.69% |
|  | Darwin | 1 | 0.345% |
|  | Downing | 3 | 1.03% |
|  | Emmanuel | 5 | 1.72% |
|  | Fitzwilliam | 2 | 0.690% |
|  | Girton | 8 | 2.76% |
|  | Gonville & Caius | 5 | 1.72% |
|  | Homerton | 4 | 1.38% |
|  | Hughes Hall | 3 | 1.03% |
|  | Jesus | 3 | 1.03% |
|  | King's | 2 | 0.690% |
|  | Lucy Cavendish | 3 | 1.03% |
|  | Magdalene | 2 | 0.690% |
|  | Murray Edwards | 6 | 2.07% |
|  | Newnham | 5 | 1.72% |
|  | Pembroke | 5 | 1.72% |
|  | Queen's | 5 | 1.72% |
|  | Robinson | 4 | 1.38% |
|  | Selwyn | 5 | 1.72% |
|  | Sidney Sussex | 2 | 0.690% |
|  | St Catharine's | 2 | 0.690% |
|  | St Edmund's | 2 | 0.690% |
|  | St John's | 3 | 1.03% |
|  | Trinity | 10 | 3.45% |
|  | Trinity Hall | 5 | 1.72% |
|  | Wolfson | 2 | 0.690% |
|  | Missing | 176 | 60.7% |
| Course | Undergraduate | 90 | 31.0% |
|  | Postgraduate | 25 | 8.62% |
|  | Missing | 175 | 60.3% |

### Appendix 3: Full results of univariate analysis

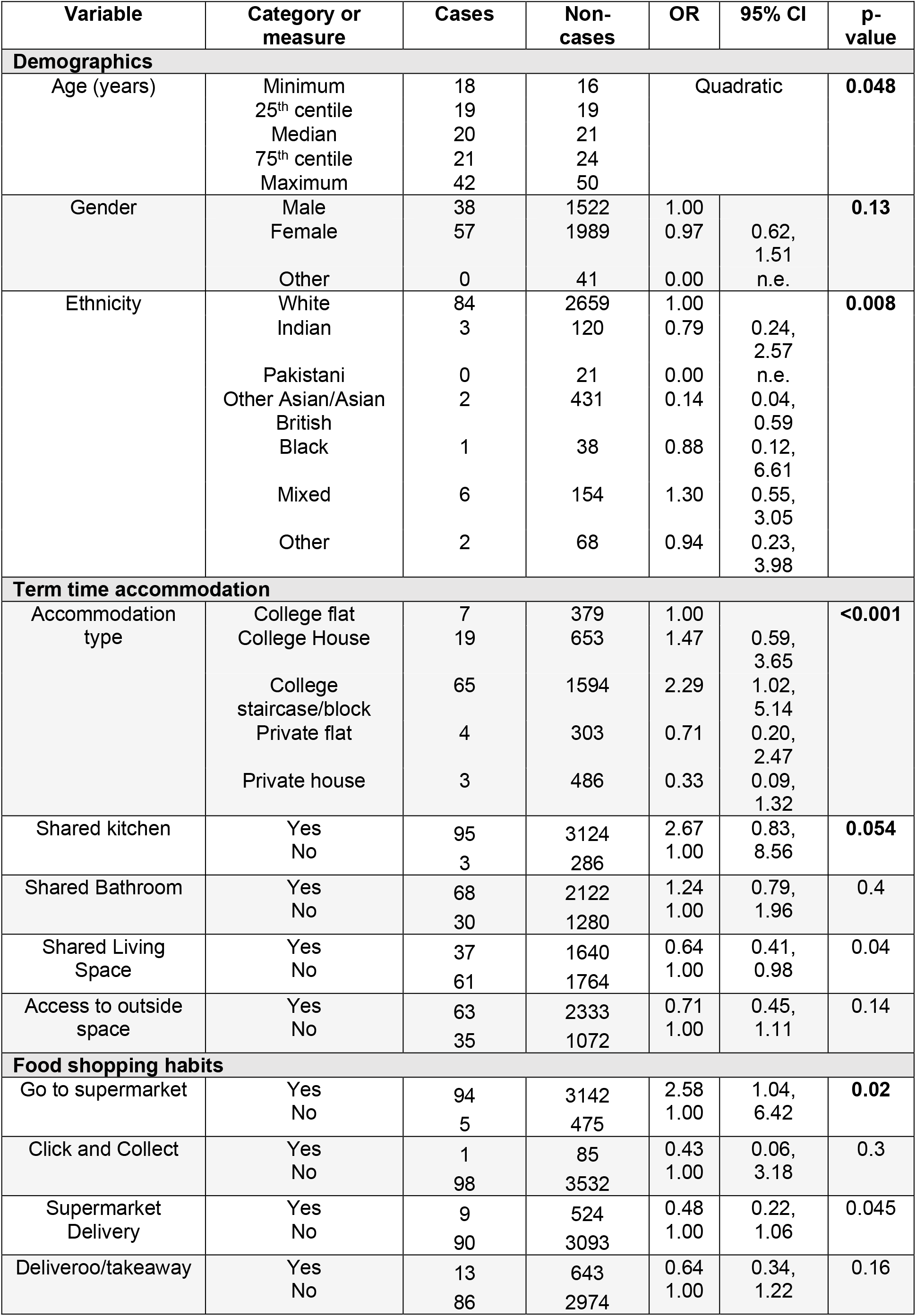
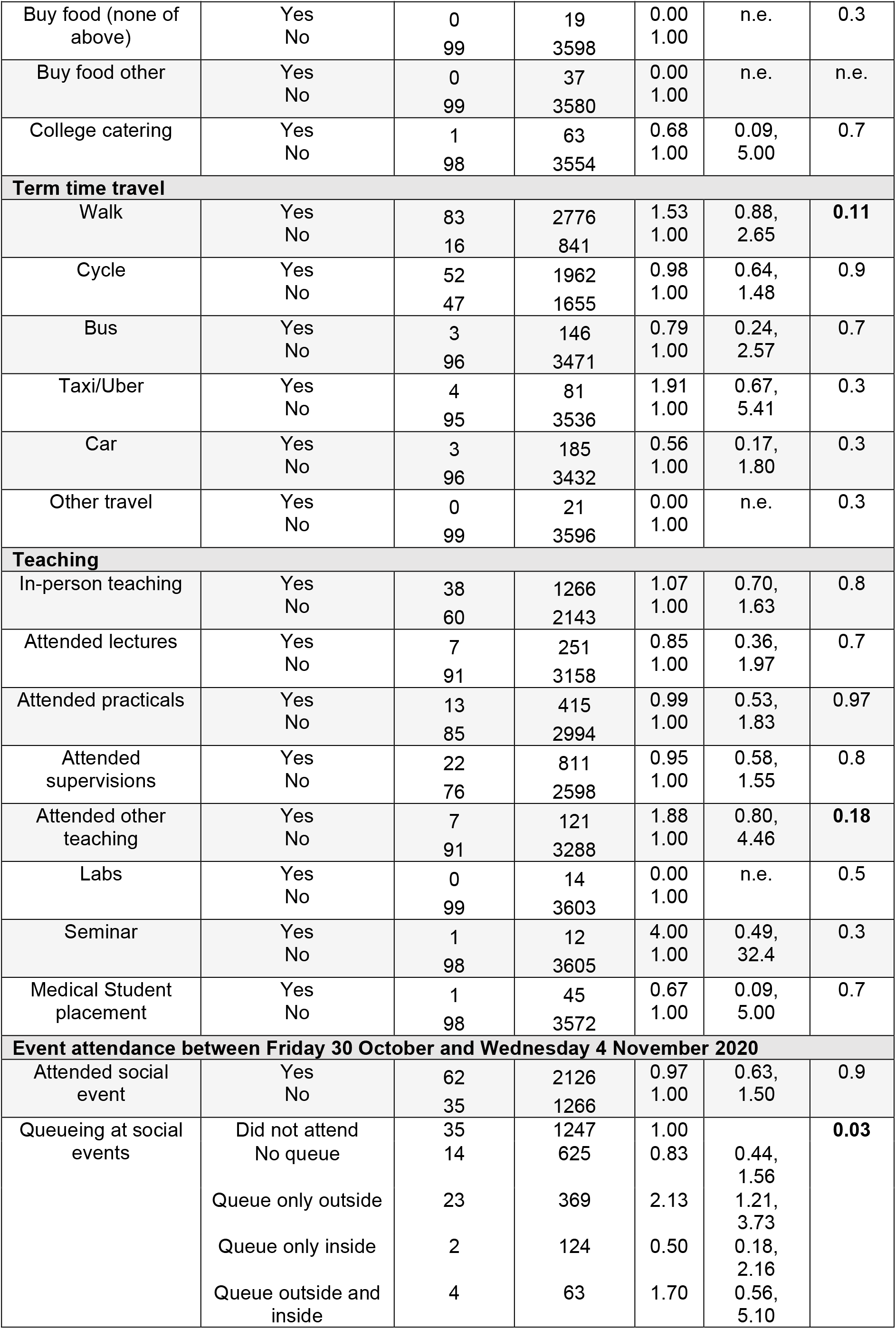
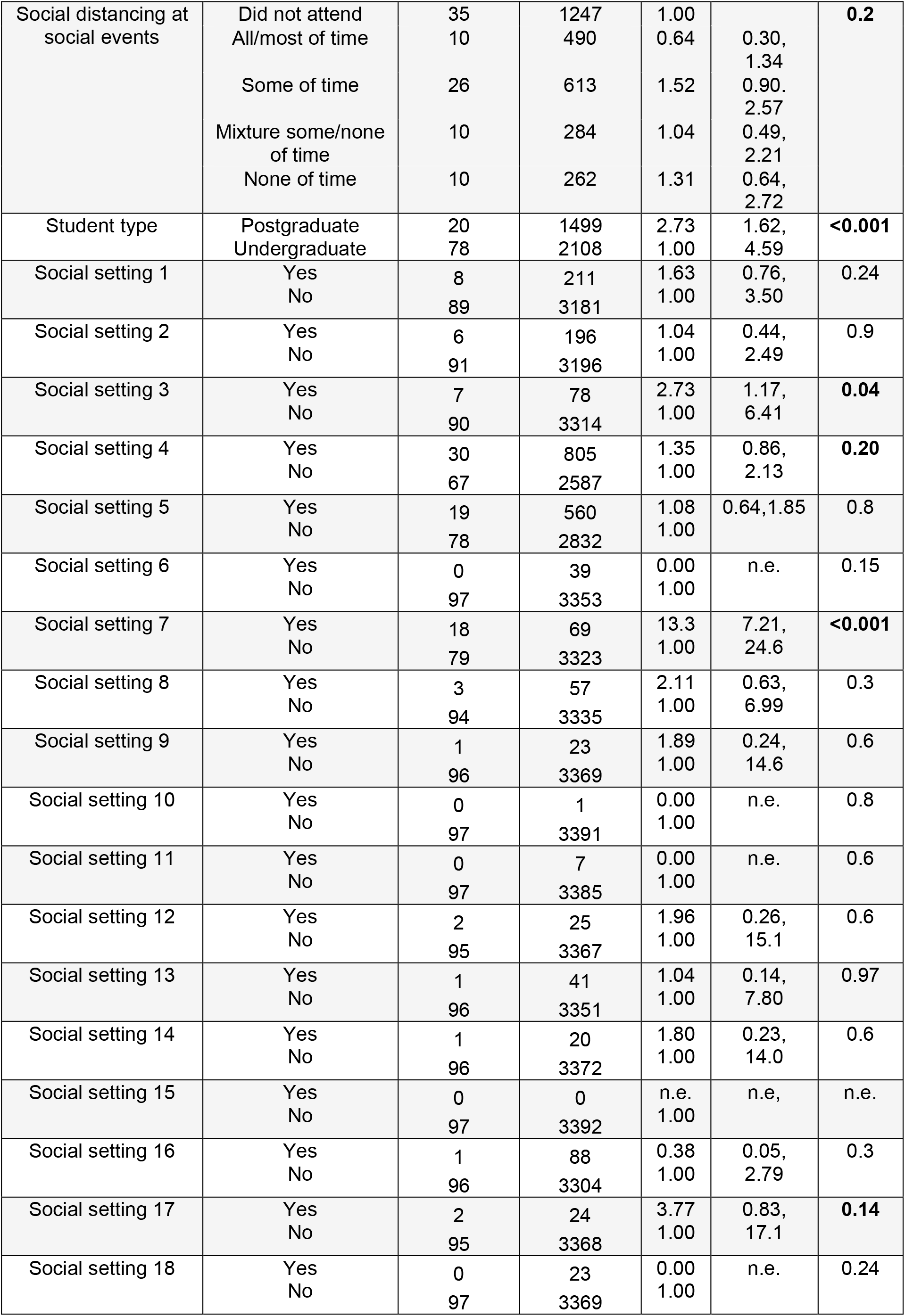
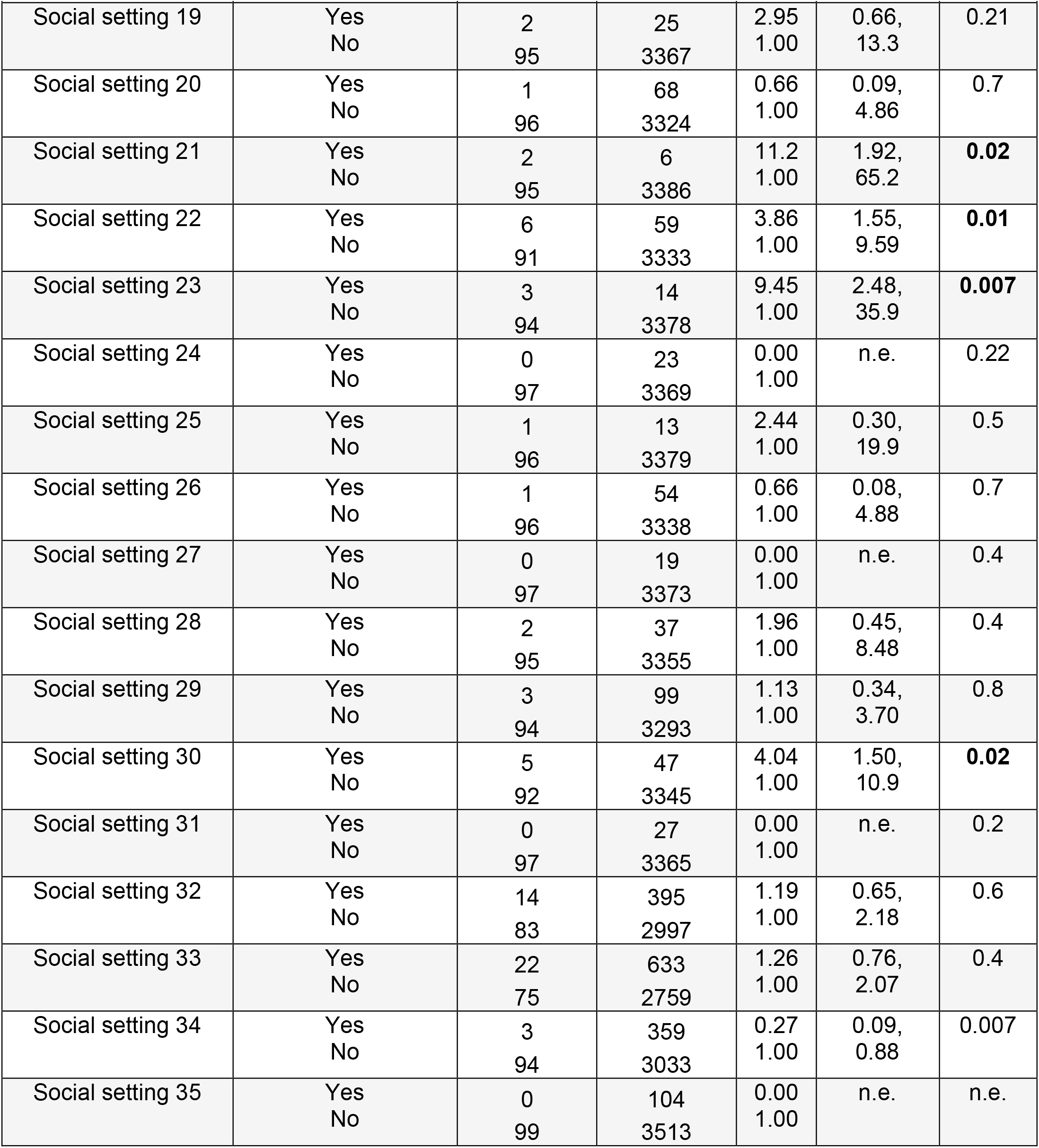

### Appendix 4: Queuing and social distancing at social settings of interest

Appendix 4: Queuing and social distancing at social settings of interest.
| Venue | Characteristic |  | Overall |  | Cases |  | Non-cases |  |
| --- | --- | --- | --- | --- | --- | --- | --- | --- |
| Social setting 7 | Queueing at social events | Queueing only outside | 63 | 1.6% | 16 | 16.2% | 47 | 1.3% |
|  |  | Queueing only inside | 1 | 0.0% | 0 | 0.0% | 1 | 0.0% |
|  |  | No queueing | 4 | 0.1% | 0 | 0.0% | 4 | 0.1% |
|  |  | Unknown queueing | 19 | 0.5% | 2 | 2.0% | 17 | 0.5% |
|  |  | Did not attend | 3410 | 85.7% | 79 | 79.8% | 3323 | 91.9% |
|  |  | Did not respond | 483 | 12.1% | 2 | 2.0% | 225 | 6.2% |
|  | Social distancing at events | All/most of time | 33 | 0.8% | 7 | 7.1% | 26 | 0.7% |
|  |  | Some of time | 38 | 1.0% | 9 | 9.1% | 29 | 0.8% |
|  |  | Mixture some/none of time | 1 | 0.0% | 0 | 0.0% | 1 | 0.0% |
|  |  | None of time | 7 | 0.2% | 2 | 2.0% | 5 | 0.1% |
|  |  | Unknown social distancing | 8 | 0.2% | 0 | 0.0% | 8 | 0.2% |
|  |  | Did not attend | 3410 | 85.7% | 79 | 79.8% | 3323 | 91.9% |
|  |  | Did not respond | 483 | 12.1% | 2 | 2.0% | 225 | 6.2% |
| Social setting 23 | Queueing at social events | Queueing only outside | 0 | 0.0% | 0 | 0.0% | 0 | 0.0% |
|  |  | Queueing only inside | 5 | 0.1% | 1 | 1.0% | 4 | 0.1% |
|  |  | No queueing | 8 | 0.2% | 2 | 2.0% | 6 | 0.2% |
|  |  | Unknown queueing | 4 | 0.1% | 0 | 0.0% | 4 | 0.1% |
|  |  | Did not attend | 3480 | 87.4% | 94 | 94.9% | 3378 | 93.4% |
|  |  | Did not respond | 483 | 12.1% | 2 | 2.0% | 225 | 6.2% |
|  | Social distancing at events | All/most of time | 8 | 0.2% | 1 | 1.0% | 7 | 0.2% |
|  |  | Some of time | 5 | 0.1% | 2 | 2.0% | 3 | 0.1% |
|  |  | Mixture some/none of time | 0 | 0.0% | 0 | 0.0% | 0 | 0.0% |
|  |  | None of time | 0 | 0.0% | 0 | 0.0% | 0 | 0.0% |
|  |  | Unknown social distancing | 4 | 0.1% | 0 | 0.0% | 4 | 0.1% |
|  |  | Did not attend | 3480 | 87.4% | 94 | 94.9% | 3378 | 93.4% |
|  |  | Did not respond | 483 | 12.1% | 2 | 2.0% | 225 | 6.2% |

## Notes

### Competing Interest Statement

The authors have declared no competing interest.

### Funding Statement

This work was completed for routine outbreak investigation purposes, and as such did not require nor receive any funding from third parties.

### Author Declarations

This protocol was subject to a review by the PHE Research Ethics and Governance Group. It was classified as an outbreak investigation undertaken as part of PHE's responsibility to respond to the COVID-19 current pandemic, and to inform the multi-agency response to the large rise in cases and future response. The study is anonymous and consent was requested to publish non-identifiable aggregate information derived from the study. As such this work fell outside the remit for ethical review and as no regulatory issues were identified the protocol was approved.

