## Supplementary: Figures for "Social risk factors for SARS-CoV-2 acquisition in University students: cross sectional survey"

**Supplementary Figure 1: Distribution of cases by specimen date in Cambridge City local authority, and by reported symptom onset date in University of Cambridge cohort (n=90).**

A)

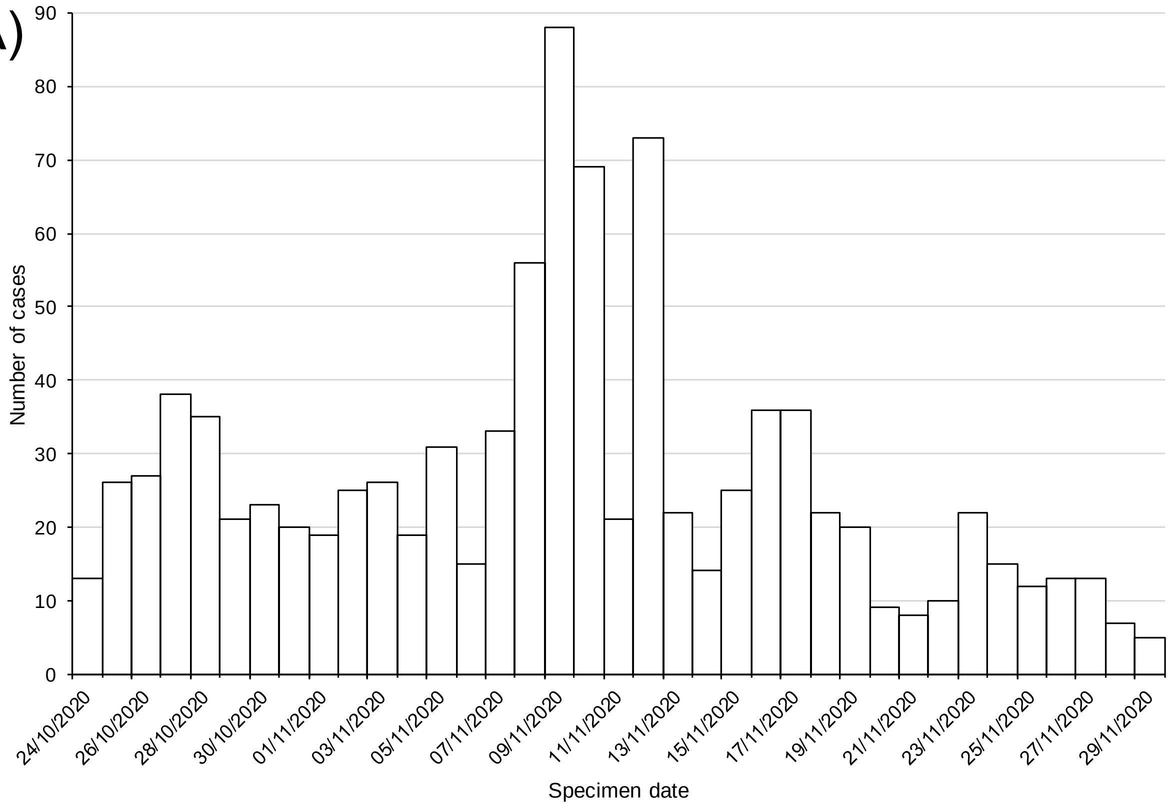


B)

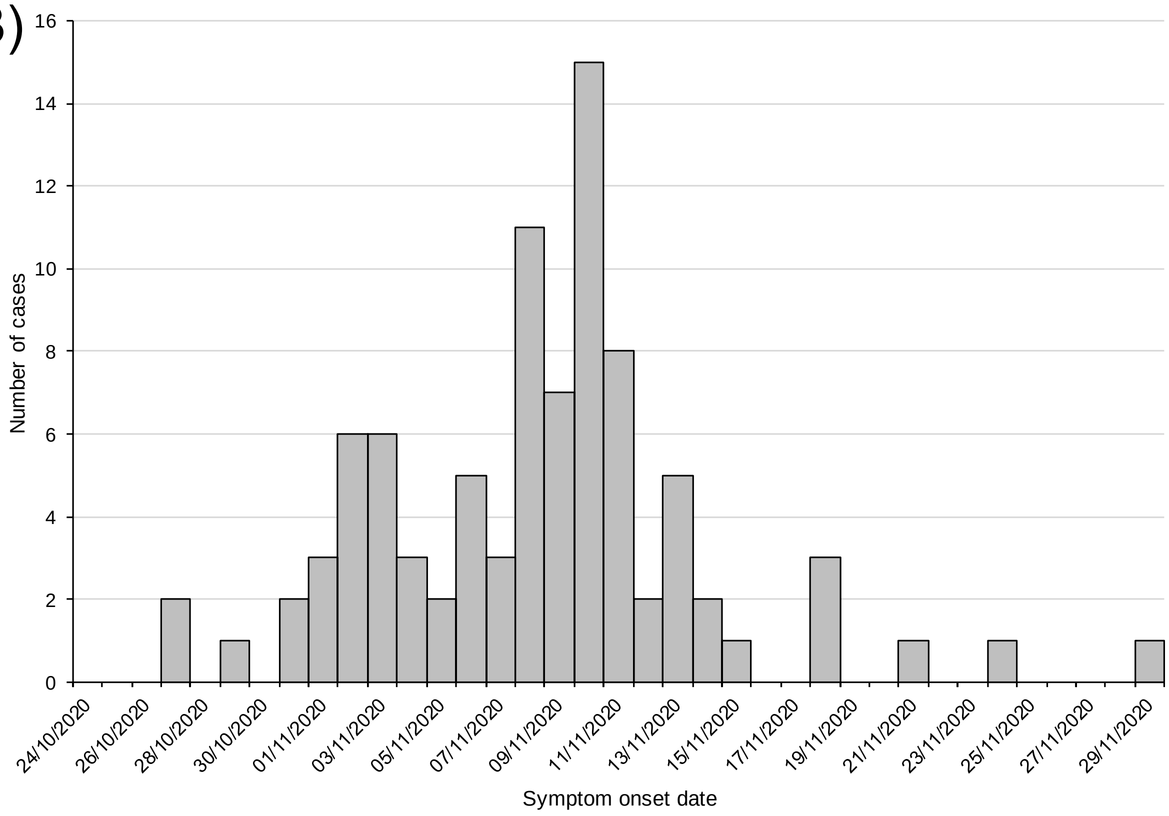
