## Supplementary: Survey for "Social risk factors for SARS-CoV-2 acquisition in University students: cross sectional survey"

Public Health England

Protecting and improving the nation's health

### Cambridge Student COVID-19 Exposure Questionnaire

Due to a number of COVID-19 outbreaks amongst students at the University of Cambridge, we are carrying out a survey so we can better understand how the virus has spread and the key exposures for students in Cambridge.

Public Health England (PHE) is an executive agency of the Department of Health and Social Care, providing the delivery of specialist public health services including investigating and controlling outbreaks of infections. We are working with the Director of Public Health for Cambridgeshire and Peterborough, Cambridge City Council, and the University of Cambridge to investigate a rise in COVID-19 cases in Cambridge City among University of Cambridge students following the week of Friday 30th October to Wednesday 4th November 2020.

To help us understand the possible sources of infection, we would be very grateful if you could take a few minutes to complete this online questionnaire. The University of Cambridge is fully supporting this investigation, allowing us to survey a defined group (cohort) so we can better understand and identify routes of exposure.

**It is very important that individuals who remained well during this time also complete this questionnaire. Your information will enable us to compare how different exposures affect the transmission of COVID-19 infection.**

**The answers you give will be kept in strict confidence, will be held in line with the General Data Protection Regulation 2018 and used solely for the purpose of this investigation, and will not be used to take action against any individuals.**

We thank you for taking part in this survey, your answers will be helping your local community to manage outbreaks in the future, and will help steer support where it is needed to break the transmission of COVID-19 in Cambridge City.

Q1 Are you a current University of Cambridge student (undergraduate or postgraduate) who has been resident in Cambridge in Michaelmas (autumn term) 2020?*

- ◯ Yes
- ◯ No

TRIAL MODE - Click here for more information

### Instructions for completing this questionnaire

**Please complete this questionnaire whether you remained well or if you became unwell. Your information will enable us to compare how different exposures affect the likelihood of COVID-19 infection.**

Please read each question carefully before you answer it and try to answer every section as far as possible. Please do not leave blanks. ‘No’ or ‘Not sure’ answers are as important as ‘Yes’ answers. If you leave a blank we cannot interpret your intended answer. There are certain questions throughout the survey that are required indicated by an asterisk (*) at the end of the question. These need to be completed before moving to the next page.

Please complete this questionnaire in one sitting and do not navigate away from the page until you have reached the end of the questions.

This questionnaire should be answered by, or for any University of Cambridge student. We need separate responses for each person, so if you are filling it in on behalf of others, please complete all answers for that person, and restart the questionnaire again to complete it for yourself.

Q2 Are you completing this questionnaire **on behalf of another person?***

- ◯ Yes
- ◯ No

If you are completing the questionnaire on behalf of another person please answer the following questions as if you were them.

TRIAL MODE - Click here for more information

### Section A: General Details

This section asks for some basic information about you.

TRIAL MODE - Click here for more information

Q3 Year of birth

--Click Here--

1970
1971
1972
1973
1974
1975
1976
1977
1978
1979
1980
1981
1982
1983
1983
1984
1985
1986
1987
1988
1989
1990
1991
1992
1993
1994
1995
1996
1997
1998
1999
2000
2001
2002
2003
2004
2005
2006
2007
2008
2009
2010
2011
2012
2013
2014
2015
2016
2017
2018
2019
2020

TRIAL MODE - Click here for more information

#### Q4 Gender

- Male
- Female
- Other
- Rather not say

#### Q5 Ethnicity

- White
- Indian (Asian or Asian British)
- Pakistani (Asian or Asian British)
- Other Asian/Asian British
- Black/African/Caribbean/Black
- British Mixed/Multiple ethnic groups
- Other ethnic group
- Rather not say

#### Q6 College

--Click Here--

- Christ's College
- Churchill College
- Clare Hall
- Corpus Christi College
- Darwin College
- Downing College
- Emmanuel College
- Fitzwilliam College
- Girton College
- Gonville & Caius College
- Homerton College
- Hughes Hall
- Jesus College
- King's College
- Lucy Cavendish College
- Magdalene College
- Murray Edwards College
- Newnham College
- Pembroke College
- Peterhouse
- Queen's College
- Robinson College
- Selwyn College
- Sidney Sussex College
- St Catherine's College
- St Edmund's College
- St John's College
- Trinity College
- Trinity Hall
- Wolfson College

TRIAL MODE - Click here for more information

Q7 Are you an undergraduate or postgraduate student?

- ◯ Undergraduate
- ◯ Postgraduate

TRIAL MODE - Click here for more information

### Section B: Clinical details

#### Illness

Q8 Have you had any of the following symptoms **since Friday 16th October 2020?***

| | Yes | No |
|---|---|---|
| Fever/high temperature (which means you feel hot to touch on your chest or back) | ◯ | ◯ |
| New continuous cough (coughing a lot for more than an hour, or 3 or more coughing episodes in 24 hours) | ◯ | ◯ |
| Loss/change of sense of taste or smell | ◯ | ◯ |
| Shortness of breath | ◯ | ◯ |
| Sore throat | ◯ | ◯ |
| Runny nose | ◯ | ◯ |
| Headache | ◯ | ◯ |
| Muscle aches | ◯ | ◯ |
| Extreme fatigue/tiredness | ◯ | ◯ |
| Diarrhoea | ◯ | ◯ |
| Vomiting | ◯ | ◯ |
| Other symptoms | ◯ | ◯ |

Q9 If you experienced other symptoms than those listed above, please provide details

TRIAL MODE - Click here for more information

### Section B: Clinical details

#### Illness continued

Q10 What date did you first start to feel unwell?

[ ]

Q11 Do you still feel unwell?

- ○ Yes
- ○ No

Q12 If you no longer have symptoms, approximately what date did they end?

[ ]

Q13 Have you had face to face contact with anyone **who was ill** with fever, new continuous cough, or loss/change of sense of taste or smell **since Friday 16th October 2020?***

- ○ Yes
- ○ No
- ○ Not sure

Q14 If yes, what was the **earliest date** of your face to face contact with this person/these people?

[ ]

TRIAL MODE - Click here for more information

#### Section B: Clinical details

##### Specimen

Q15 Have you supplied a specimen for COVID-19 testing **since Friday 30th October?***

- Yes
- No
- Not sure

Q16 If yes, was this through University of Cambridge-organised testing or through other routes such as NHS testing?

- University of Cambridge Screening Programme
- Cambridge University Hospital screening programme
- NHS testing
- Other
- Not sure

If other please specify

Q17 If yes, have you had a **positive individual** test result **since Friday 30th October?**

- Yes
- No
- Still waiting for the result
- Not sure

Q18 If you had a **positive individual** test result, when did you submit that sample?

Q19 If you have had a **positive individual** test result **since Friday 30th October**, was this in relation to any **illness** including fever, new continuous cough, or loss/change of sense of taste or smell?

- Yes
- No

Q20 If you have **not** had a positive individual test result, what test results have you had **since Friday 30th October?**

| | Yes | No |
|---|---|---|
| Positive pooled test sample | ◯ | ◯ |
| Negative pooled test sample | ◯ | ◯ |
| Negative individual test sample | ◯ | ◯ |

TRIAL MODE - Click here for more information

Q21 If you have had a pooled positive test sample, what was the date of your **most recent** pooled positive test result?

[ ]

Q22 If you have had negative test results, what was the date of your **most recent** test result?

[ ]

Q23 Is there any additional relevant information you think might be useful to us about your COVID-19 tests?

[ ]

#### Medical Consultation

Q24 Did you consult your College Nurse, 111, your GP or another doctor, or visit A&E as a result of this illness?

- ○ Yes
- ○ No

Q25 If yes, what kind of medical advice did you consult?

- ☐ College Nurse
- ☐ NHS 111
- ☐ GP or another doctor
- ☐ Visit A&E
- ☐ Other

If other please specify

[ ]

Q26 Were you admitted to hospital for more than four hours as a result of this illness?

- ○ Yes
- ○ No

Q27 Hospital name

[ ]

Q28 What date were you admitted to hospital as a result of this illness?

[ ]

TRIAL MODE - Click here for more information

Q29 Are you still in hospital?

- ◯ Yes
- ◯ No

Q30 If no, what date were you discharged from hospital?

TRIAL MODE - Click here for more information

### Section C: Other exposures

#### Accomodation

Q31 What type of accomodation best describes where you live during term time?

- ◯ College staircase/block
- ◯ College flat
- ◯ College house
- ◯ Private flat
- ◯ Private house

Q32 Do you share a kitchen/kitchenette with other people in your term time accommodation?

- ◯ Yes
- ◯ No

Q33 Do you share a bathroom with other people in your term time accommodation?

- ◯ Yes
- ◯ No

Q34 Do you have a shared living space (e.g. living room, dining room) with other people in your term time accommodation?

- ◯ Yes
- ◯ No

Q35 Do you have access to outside areas in your term time accommodation (i.e. even when isolating)?

- ◯ Yes
- ◯ No

#### Teaching

Q36 Did you attend any in-person teaching between Friday 30th October and Wednesday 4th November 2020?

- ◯ Yes
- ◯ No

Q37 If yes, when did you attend in-person teaching and what best defines the type of teaching?

| | Friday 30th October | Saturday 31st October | Sunday 1st November | Monday 2nd November | Tuesday 3rd November | Wednesday 4th November |
|---|---|---|---|---|---|---|
| Lecture | ☐ | ☐ | ☐ | ☐ | ☐ | ☐ |
| Practicals | ☐ | ☐ | ☐ | ☐ | ☐ | ☐ |
| Supervisions/tutorials/one to one meetings | ☐ | ☐ | ☐ | ☐ | ☐ | ☐ |

TRIAL MODE - Click here for more information

Other ☐ ☐ ☐ ☐ ☐ ☐

Q38 If you attended other kinds of in-person teaching, please provide details below:

#### Food shopping

Q39 How do you usually buy food?

- ☐ Go to supermarket
- ☐ Click and collect
- ☐ Supermarket delivery to residence
- ☐ Deliveroo or takeaway
- ☐ None of the above
- ☐ Other

If other please specify

Q40 If you went to a supermarket, please describe which supermarket(s) you visited below (i.e. chain, location, frequency)?

#### Travel

Q41 How do you usually travel during term time?

- ☐ Walk
- ☐ Cycle
- ☐ Bus
- ☐ Taxi or Uber
- ☐ Car
- ☐ Other

Q42 In the **two weeks prior** to Friday 30th October 2020 (Friday 16th October to Thursday 29th October), did you travel outside the UK?*

- ○ Yes
- ○ No

TRIAL MODE - Click here for more information

Q43 If yes, can you please provide details of this travel (i.e. country visited, dates)

TRIAL MODE - Click here for more information

#### Section D: Event attendance

The following is a list of events and venues that you may have attended prior to the national lockdown on Thursday 5th November 2020.

To help us identify potential sources for the recent increase in COVID-19 cases across the University of Cambridge, we would be grateful if you could check one box for each of the events and venues listed below, indicating whether you attended or not.

**The information you provide to us will be held in strict confidence, and will not be used to take action against any individuals.**

Q44 Did you go out socially or meet with friends between Friday 30th October and Wednesday 4th November 2020?*

- ◯ Yes
- ◯ No

TRIAL MODE - Click here for more information

#### Section D: Event attendance

Q45 If yes, when and where did you attend formal or informal social events?

| | Friday 30th October | Saturday 31st October | Sunday 1st Nove mber | Monday 2nd Nov ember | Tuesday 3rd Nove mber | Wednesd ay 4th N ovember | Did not attend |
|---|---|---|---|---|---|---|---|
| SS1 | ☐ | ☐ | ☐ | ☐ | ☐ | ☐ | ☐ |
| SS2 | ☐ | ☐ | ☐ | ☐ | ☐ | ☐ | ☐ |
| SS3 | ☐ | ☐ | ☐ | ☐ | ☐ | ☐ | ☐ |
| SS4 | ☐ | ☐ | ☐ | ☐ | ☐ | ☐ | ☐ |
| SS5 | ☐ | ☐ | ☐ | ☐ | ☐ | ☐ | ☐ |
| SS6 | ☐ | ☐ | ☐ | ☐ | ☐ | ☐ | ☐ |
| SS7 | ☐ | ☐ | ☐ | ☐ | ☐ | ☐ | ☐ |
| SS8 | ☐ | ☐ | ☐ | ☐ | ☐ | ☐ | ☐ |
| SS9 | ☐ | ☐ | ☐ | ☐ | ☐ | ☐ | ☐ |
| SS10 | ☐ | ☐ | ☐ | ☐ | ☐ | ☐ | ☐ |
| SS11 | ☐ | ☐ | ☐ | ☐ | ☐ | ☐ | ☐ |
| SS12 | ☐ | ☐ | ☐ | ☐ | ☐ | ☐ | ☐ |
| SS13 | ☐ | ☐ | ☐ | ☐ | ☐ | ☐ | ☐ |
| SS14 | ☐ | ☐ | ☐ | ☐ | ☐ | ☐ | ☐ |
| SS15 | ☐ | ☐ | ☐ | ☐ | ☐ | ☐ | ☐ |
| SS16 | ☐ | ☐ | ☐ | ☐ | ☐ | ☐ | ☐ |
| SS17 | ☐ | ☐ | ☐ | ☐ | ☐ | ☐ | ☐ |
| SS18 | ☐ | ☐ | ☐ | ☐ | ☐ | ☐ | ☐ |
| SS19 | ☐ | ☐ | ☐ | ☐ | ☐ | ☐ | ☐ |
| SS20 | ☐ | ☐ | ☐ | ☐ | ☐ | ☐ | ☐ |
| SS21 | ☐ | ☐ | ☐ | ☐ | ☐ | ☐ | ☐ |
| SS22 | ☐ | ☐ | ☐ | ☐ | ☐ | ☐ | ☐ |
| SS23 | ☐ | ☐ | ☐ | ☐ | ☐ | ☐ | ☐ |
| SS24 | ☐ | ☐ | ☐ | ☐ | ☐ | ☐ | ☐ |
| SS25 | ☐ | ☐ | ☐ | ☐ | ☐ | ☐ | ☐ |
| SS26 | ☐ | ☐ | ☐ | ☐ | ☐ | ☐ | ☐ |
| SS27 | ☐ | ☐ | ☐ | ☐ | ☐ | ☐ | ☐ |
| SS28 | ☐ | ☐ | ☐ | ☐ | ☐ | ☐ | ☐ |
| SS29 | ☐ | ☐ | ☐ | ☐ | ☐ | ☐ | ☐ |

TRIAL MODE - Click here for more information

| | | | | | | | |
|---|---|---|---|---|---|---|---|
| SS30 | ☐ | ☐ | ☐ | ☐ | ☐ | ☐ | ☐ |
| SS31 | ☐ | ☐ | ☐ | ☐ | ☐ | ☐ | ☐ |
| SS32 | ☐ | ☐ | ☐ | ☐ | ☐ | ☐ | ☐ |
| SS33 | ☐ | ☐ | ☐ | ☐ | ☐ | ☐ | ☐ |
| SS34 | ☐ | ☐ | ☐ | ☐ | ☐ | ☐ | ☐ |

Q46 If other, please provide details of the event/venue below (i.e. location, description, approximate number of people there).

Q47 If yes, what was your experience like at that event/venue?

| | Queued outdoors | Queued indoors | No queueing | 2m social distancing all/most of the time | 2m social distancing some of the time | 2m social disal distancing little/none of the time |
|---|---|---|---|---|---|---|
| SS1 | ☐ | ☐ | ☐ | ☐ | ☐ | ☐ |
| SS2 | ☐ | ☐ | ☐ | ☐ | ☐ | ☐ |
| SS3 | ☐ | ☐ | ☐ | ☐ | ☐ | ☐ |
| SS4 | ☐ | ☐ | ☐ | ☐ | ☐ | ☐ |
| SS5 | ☐ | ☐ | ☐ | ☐ | ☐ | ☐ |
| SS6 | ☐ | ☐ | ☐ | ☐ | ☐ | ☐ |
| SS7 | ☐ | ☐ | ☐ | ☐ | ☐ | ☐ |
| SS8 | ☐ | ☐ | ☐ | ☐ | ☐ | ☐ |
| SS9 | ☐ | ☐ | ☐ | ☐ | ☐ | ☐ |
| SS10 | ☐ | ☐ | ☐ | ☐ | ☐ | ☐ |
| SS11 | ☐ | ☐ | ☐ | ☐ | ☐ | ☐ |
| SS12 | ☐ | ☐ | ☐ | ☐ | ☐ | ☐ |
| SS13 | ☐ | ☐ | ☐ | ☐ | ☐ | ☐ |
| SS14 | ☐ | ☐ | ☐ | ☐ | ☐ | ☐ |
| SS15 | ☐ | ☐ | ☐ | ☐ | ☐ | ☐ |
| SS16 | ☐ | ☐ | ☐ | ☐ | ☐ | ☐ |
| SS17 | ☐ | ☐ | ☐ | ☐ | ☐ | ☐ |
| SS18 | ☐ | ☐ | ☐ | ☐ | ☐ | ☐ |

TRIAL MODE - Click here for more information

| | | | | | | |
|---|---|---|---|---|---|---|
| SS19 | ☐ | ☐ | ☐ | ☐ | ☐ | ☐ |
| SS20 | ☐ | ☐ | ☐ | ☐ | ☐ | ☐ |
| SS21 | ☐ | ☐ | ☐ | ☐ | ☐ | ☐ |
| SS22 | ☐ | ☐ | ☐ | ☐ | ☐ | ☐ |
| SS23 | ☐ | ☐ | ☐ | ☐ | ☐ | ☐ |
| SS24 | ☐ | ☐ | ☐ | ☐ | ☐ | ☐ |
| SS25 | ☐ | ☐ | ☐ | ☐ | ☐ | ☐ |
| SS26 | ☐ | ☐ | ☐ | ☐ | ☐ | ☐ |
| SS27 | ☐ | ☐ | ☐ | ☐ | ☐ | ☐ |
| SS28 | ☐ | ☐ | ☐ | ☐ | ☐ | ☐ |
| SS29 | ☐ | ☐ | ☐ | ☐ | ☐ | ☐ |
| SS30 | ☐ | ☐ | ☐ | ☐ | ☐ | ☐ |
| SS31 | ☐ | ☐ | ☐ | ☐ | ☐ | ☐ |
| SS32 | ☐ | ☐ | ☐ | ☐ | ☐ | ☐ |
| SS33 | ☐ | ☐ | ☐ | ☐ | ☐ | ☐ |
| SS34 | ☐ | ☐ | ☐ | ☐ | ☐ | ☐ |

TRIAL MODE - Click here for more information

#### Section E: Additional Information

Q48 Please give any additional relevant information you think might be useful to us, including comments about your illness, your accommodation, or the events/venues you attended between Friday 30th October and Wednesday 4th November.

TRIAL MODE - Click here for more information

### Section F: Additional Information

At present your responses are not linked to you as an individual.

PHE are involved in a number of research projects understanding the spread of COVID-19 in universities, including genomic analysis of positive test samples.

Are you happy to be contacted by the PHE investigation team to seek your consent for involvement in further research? Full details of the research and how your data would be used will be provided at that time.

If so, please enter your email address below.

Q49 Email address

[ ]

TRIAL MODE - Click here for more information

Thank you for taking the time to complete this survey. Please hit submit where you will be taken to a finishing page, and your responses saved.

TRIAL MODE - Click here for more information
